# Similar Cardiac and Neurological Outcomes of Chest Compression-Only Versus Standard CPR but lower survival with CPR after sensitivity analysis. : A Meta-analysis of Randomized Controlled Trials and Observational Cohorts in Out-of-Hospital Cardiac Arrest (OHCA)

**DOI:** 10.1101/2025.08.04.25332748

**Authors:** Ashkan Bahrami, Reza Eshraghi, Ehsan Amini-Salehi, Bahar Darouei, Reza Amani-Beni, Mazaheri-Tehrani Sadegh, Sina Sadati, Pouya Ebrahimi, Mohammad Reza Movahed

## Abstract

**Background:** Out-of-hospital cardiac arrest (OHCA) has high mortality, and bystander cardiopulmonary resuscitation (CPR) improves outcomes. The effectiveness of chest compression-only CPR (CCO) versus standard CPR (sCPR) with ventilation remains unclear. This meta-analysis assesses their impact on survival, neurological recovery, and return of spontaneous circulation (ROSC).

**Methods:** A systematic review and meta-analysis followed PRISMA guidelines. PubMed, Scopus, Web of Science, Embase, Google Scholar, and Cochrane Library were searched. Eligible studies included RCTs and observational studies on adult OHCA. Pediatric and non-original studies were excluded. Primary outcomes were survival to hospital discharge (SHD) and neurological recovery, while secondary outcomes included ROSC, survival to hospital admission, and 24-hour and one-month survival. Study quality was assessed using ROB-2 for RCTs and the Newcastle-Ottawa Scale (NOS) for observational studies. A random-effects model was applied, and publication bias was evaluated.

**Results:** A total of 18 studies (5 RCTs, 13 observational) with 232,655 OHCA cases were analyzed. SHD rates showed no significant difference (OR = 0.85, 95% CI: 0.61–1.19, P = 0.29). Favorable neurological outcomes were similar (OR = 0.87, 95% CI: 0.64–1.20, P = 0.32). Prehospital ROSC rates were comparable (OR = 1.06, 95% CI: 0.89–1.27, P = 0.43). No difference was found in survival to hospital admission (OR = 1.12, 95% CI: 0.53– 2.29, P = 0.34). For 24-hour mortality, no difference was found (OR = 0.92, 95% CI: 0.83–1.01, P = 0.07), but sCPR had lower survival after sensitivity analysis (OR = 0.90, 95% CI: 0.82–1.00, P = 0.04). One-month mortality was similar (OR = 1.26, 95% CI: 0.98–1.62, P = 0.07), but sCPR had a higher risk after outlier removal (OR = 1.32, 95% CI: 1.02–1.71, P = 0.03). Hospital discharge rates showed no difference (OR = 0.79, 95% CI: 0.36–1.72, P = 0.55). SHD with favorable neurological outcomes did not differ significantly (OR = 1.47, 95% CI: 0.09–22.68, P = 0.61). Subgroup analyses indicated sCPR benefits in witnessed arrests and shockable rhythms.

**Conclusion:** No significant differences were found between CCO and sCPR in key survival outcomes. While sCPR may benefit specific subgroups, CCO remains an effective alternative that increases bystander participation. Further research is needed to refine guidelines

## Introduction

Out-of-hospital cardiac arrest (OHCA) has been defined as the stoppage of the heart’s mechanical action outside of the hospital (2, 1). The global prevalence of OHCA is increasing, with the most recent estimates indicating around 3.8 million OHCA episodes per year (3, 4). Considering these developments, the survival and prognosis, and rates for OHCA have not improved over time (5). The majority of unpredictable OHCAs are witnessed by the general public. Bystander cardiopulmonary resuscitation (CPR) may improve outcomes for OHCA (6–10). In the study by Nadolny et al., 35.1% of OHCA patients recover spontaneously, while only 28.7% received hospitalization (11). European Resuscitation Council (ERC) and the American Heart Association (AHA) promote excellence in quality CPR (12, 13), The 2020 guidelines issued by the AHA stressed that unprofessional bystanders need to start CPR for OHCAs immediately (12). Basic life support (BLS) or CPR or may increase survival by up to three times (14). Early CPR may minimize mortality after an out-of-hospital cardiac arrest and enhance neurological performance (15–17). Regardless of the benefits, bystander-initiated CPR is frequently greeted with reluctance and delay due to fears of exacerbating the situation, insufficient skill, and concerns regarding mouth-to-mouth ventilation (18, 19). Additionally, there are presently two methods for doing bystander CPR in OHCA: standard CPR (S-CPR) and chest compression-only CPR (CCO). The initial approach is the conventional way of doing cycles that consists of 30 compressions followed by two ventilations. The second method is focused on continuous chest compression with no breaks for breathing rescue, to encourage people to engage in more frequent attempted resuscitation (20). CO-CPR may produce many more compressions each minute and is simpler for bystanders to learn and practice than sCPR (21). Nevertheless, the outcomes of bystanders performing CO-CPR remain rather disputed. Prior researches indicate that CO-CPR may contribute to fatigue and decreased compression depth, limiting its efficacy(22) A meta-analysis research found no significant variations in resuscitation outcomes between sCPR and CCO for OHCA (23–25). On the other hand, a European study found that when bystanders give ventilations in addition to chest compressions, patients have a greater chance of surviving until discharge (26, 27).

Despite the benefits of bystander CPR, the optimal approach CCO or sCPR remains debated. CCO is easier to perform and increases bystander participation but may lead to inadequate oxygenation. In contrast, sCPR includes ventilation but is associated with reluctance to perform mouth-to-mouth. Prior meta-analyses have shown conflicting results, with some reporting no significant differences, while others suggest sCPR may improve survival. Most studies focus on either randomized controlled trials (RCTs) or observational data separately, without integrating both designs. Additionally, neurological outcomes remain underexplored. This study conducts a comprehensive meta-analysis of RCTs and observational studies to compare CCO and sCPR in OHCA, assessing survival, neurological recovery, and ROSC to provide clearer guidance for CPR strategies and future resuscitation guidelines.

## Methods

### Study Design and Protocol Registration

This meta-analysis was conducted following the Preferred Reporting Items for Systematic Reviews and Meta-Analyses (PRISMA) (28)(*SUPPLEMENTARY Table 1. & 2)*, and also a systematic literature search, data extraction, quality assessment, and statistical analyses were performed to evaluate the comparative effectiveness of CCO versus sCPR with ventilation for adults experiencing OHCA.

### Search Strategy and Data Sources

A systematic search was conducted across PubMed, Scopus, Web of Science, Embase, Google Scholar, and Cochrane Library using MeSH terms, Emtree terms, and free-text keywords with Boolean operators (AND, OR). The search included terms related to CPR techniques ("resuscitation" OR "CPR" OR "chest compression-only" OR "conventional CPR") AND OHCA conditions ("cardiac arrest" OR "heart attack" OR "sudden cardiac death" OR "VF" OR "VT" OR "PEA" OR "asystole") AND study design filters ("Randomized Controlled Trials" OR "controlled clinical trials" OR "double-blind method") AND outcome measures ("mortality" OR "survival" OR "neurological outcomes" OR "quality of life" OR "spontaneous circulation"). The search strategy followed the PICO framework, ensuring the inclusion of all relevant studies evaluating CCO versus sCPR. The full search strategy is available in Supplementary Materials *(SUPPLEMENTARY Table 3.*). No language restrictions were applied during the initial search, but only English-language full-text studies were included in the final selection.

### Eligibility Criteria

Studies were selected based on the following inclusion and exclusion criteria.

### Inclusion Criteria

Original studies, including randomized controlled trials (RCTs) and observational cohort studies, with direct comparisons between CCO and sCPR were included. Studies with adult populations (≥18 years old) who experienced OHCA of presumed cardiac origin were considered. The intervention group consisted of patients whom CCO was performed by bystanders, while the comparator group consisted of those who received sCPR, including both chest compressions and ventilation. Studies were included if they reported at least one of the primary or secondary outcomes relevant to the analysis.

### Exclusion Criteria

Studies focusing on pediatric patients (<18 years old) were excluded due to differences in AHA/ACC resuscitation guidelines(29–33). Studies that investigated only one arm (CCO or sCPR) without a direct comparison between the two techniques were not included. Non-original studies, including systematic reviews, meta-analyses, case reports, case series, and conference abstracts, were excluded. Studies conducted in settings other than OHCA, such as in-hospital cardiac arrests or drowning-related arrests, were not included. Studies with incomplete or non-separable data for adult OHCA populations and non-English full-text articles were also excluded.

### Study Selection and Data Extraction

The study selection process followed the PRISMA flow diagram (Figure 2.), with two independent reviewers, B.D. and R.A., screening titles and abstracts, followed by full-text assessments for eligible studies. Any disagreements were resolved by a third reviewer, R.E. A standardized data extraction form was used to collect study details, including author, year, country, study design, and sample size, as well as patient demographics such as age, sex, location of arrest (home, outdoor, indoor, public places), and witnessed status. Arrest characteristics, including the first recorded rhythm (VF/VT, PEA, asystole), EMS response time, and the use of AEDs, were also documented. Intervention details were extracted, specifying whether CPR was CCO or sCPR, and whether dispatcher-assisted CPR was involved. The cardiac outcomes assessed included SHD, ROSC, survival to hospital admission after ROSC, 24-hour survival, and one-month survival. These outcomes were analyzed to determine whether CCO or sCPR resulted in better overall survival rates across different timeframes. The neurological outcomes were evaluated at multiple times to assess the impact of CCO and sCPR on brain function and recovery. These included survival to hospital discharge with a favorable neurological outcome, defined as a Cerebral Performance Category (CPC) score of 1 or 2, (34). as well as overall favorable neurological outcomes across all periods and additional follow-up assessments beyond hospital discharge. Data extraction was conducted independently by two reviewers, B.D. and R.A., with any discrepancies resolved by a third reviewer, R.E. This process ensured a comprehensive evaluation of both cardiac and neurological outcomes, integrating multiple timeframes to provide a thorough comparison of CCO and sCPR in OHCA cases.

**Figure 1.**
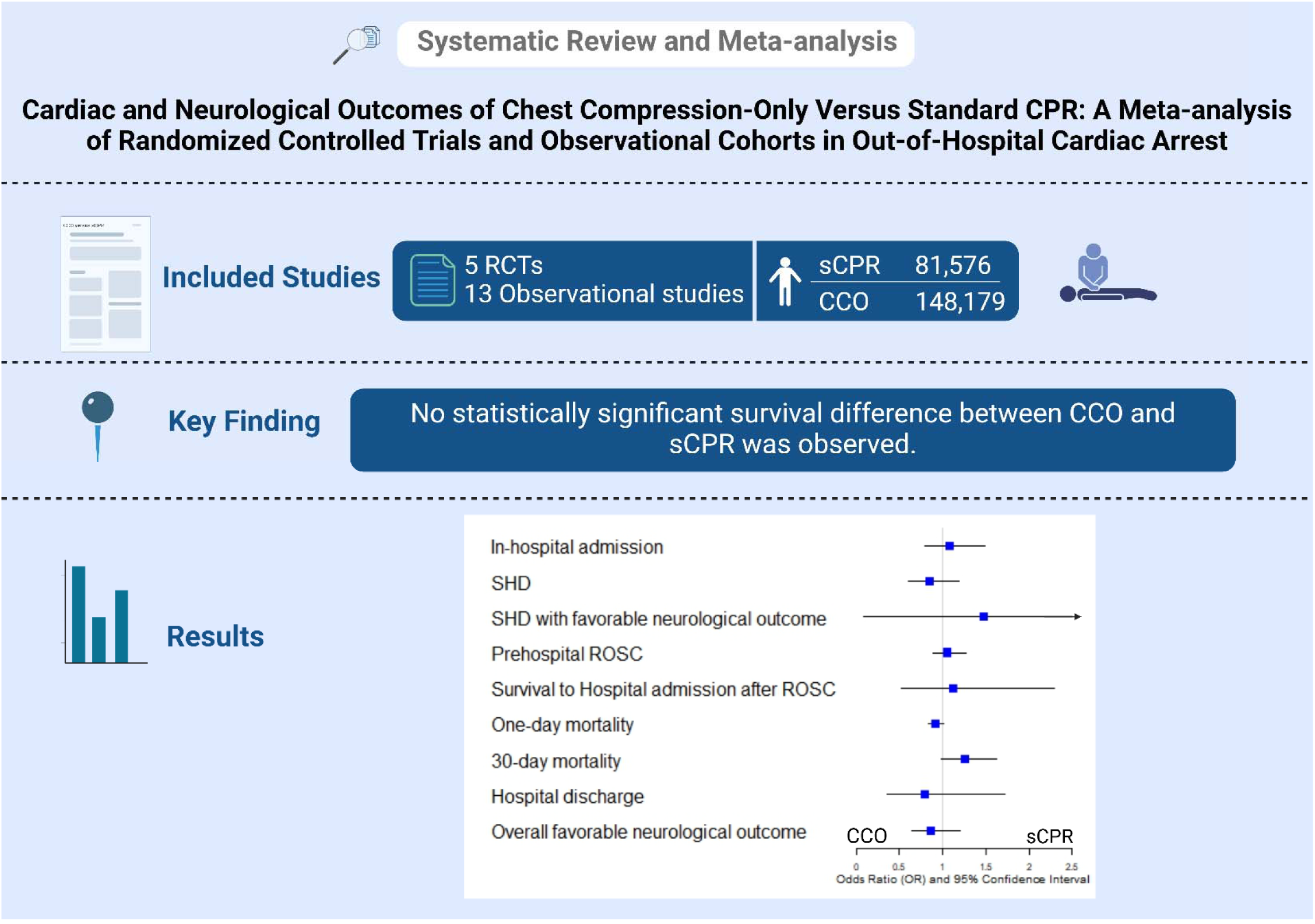
Comparative Effectiveness of Chest Compression-Only (CCO) vs. Standard CPR (sCPR) in Out-of-Hospital Cardiac Arrest (OHCA) This graphic abstract provides a visual summary of the study’s meta-analysis comparing CCO and sCPR in terms of key survival outcomes, neurological recovery, and return of spontaneous circulation (ROSC) for OHCA patients. Abbreviations: CCO, chest compression-only; sCPR, standard cardiopulmonary resuscitation (compression + ventilation); OHCA, out-of-hospital cardiac arrest; ROSC, return of spontaneous circulation.

**Figure 2.**
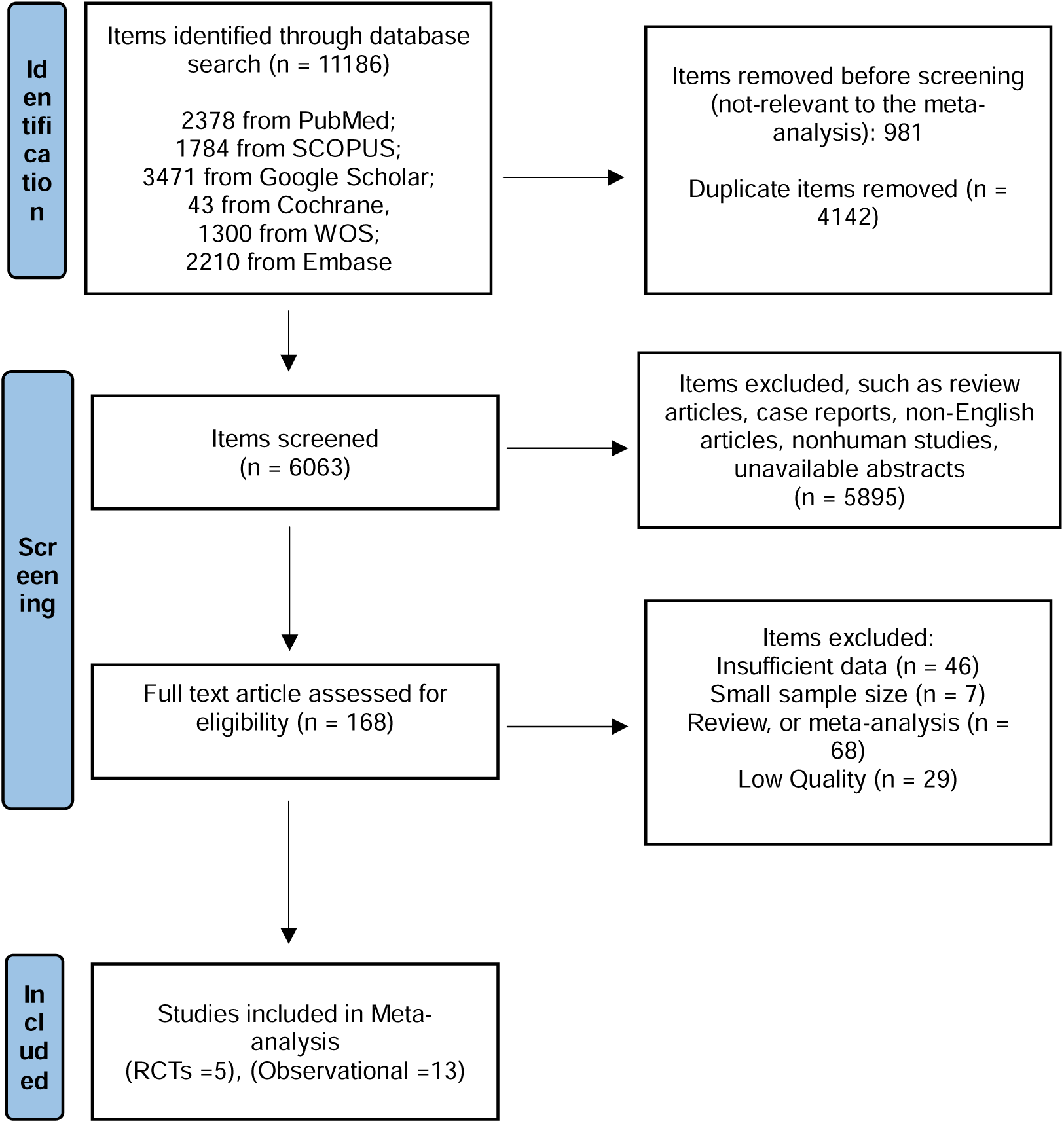
Study flow diagram.

### Quality Assessment and Risk of Bias Evaluation

The quality of included studies was independently assessed by two reviewers, S.S, and S.M. The Revised Cochrane Risk-of-Bias Tool (ROB-2) was used to assess randomized controlled trials(35), while observational studies were evaluated using the Newcastle-Ottawa Scale (NOS)(36).

### Statistical Analysis

Meta-analysis was conducted using random-effects models due to expected heterogeneity across studies. Statistical significance was set at p < 0.05, and 95% confidence intervals (CI) were reported. Pooled odds ratios (OR) with 95% CIs were calculated for each outcome. Heterogeneity was assessed using I² statistics, with I² > 50% indicating moderate to high heterogeneity. A random-effects model was used to account for between-study variability. Sensitivity analysis was conducted using the Galbraith plot to identify potential outliers, a cumulative enhancement funnel plot for examining publication bias, and the trim-and-fill method to adjust for missing studies. Publication bias was assessed using Begg’s test (38), and Egger’s test to detect small-study effects(39). Statistical power was calculated for major outcomes to determine the likelihood of detecting a true effect. Subgroup analyses were performed based on witnessed versus non-witnessed arrests, initial cardiac rhythm (shockable versus non-shockable), location of arrest, EMS response time, and AED use. All statistical analyses were performed using STATA version 18 software.

### Definitions of Key Outcomes

Return of Spontaneous Circulation (ROSC) was defined as the restoration of a palpable pulse before hospital arrival. Survival to Hospital Discharge (SHD) was defined as the patient being discharged alive from the hospital. Favorable Neurological Outcome was defined as a Cerebral Performance Category (CPC) score of 1 or 2, indicating good cerebral performance or moderate disability. EMS Response Time was defined as the time interval from call receipt to EMS arrival at the scene.

## Result

**Table 1.** Summary of the included randomized controlled trials (RCTs) and observational cohort studies comparing chest compression-only (CCO) and standard CPR (sCPR) in out-of-hospital cardiac arrest (OHCA) cases. The table presents study design, country, total sample size, and survival outcomes. One-day survival rate, 30-day survival rate. Abbreviations: CCO-CPR, chest compression-only cardiopulmonary resuscitation; sCPR, standard cardiopulmonary resuscitation (compression + ventilation); OHCA, out-of-hospital cardiac arrest; RCT, randomized controlled trial.

| Authors | YOP | Country | Design | Total | CPR |  | CCO |  |
| --- | --- | --- | --- | --- | --- | --- | --- | --- |
|  |  |  |  |  | Total | Survival | Total | Survival |
| Riva et al. (40) | 2024 | Sweden | RCT | 1250 | 610 | 168 (27.5%)<sup>a</sup> | 640 | 179 (27.9%)<sup>a</sup> |
| Svensson et al. (41) | 2010 | Sweden | RCT | 1276 | 656 | 136<sup>a</sup> (20.7%), 46 (7.01%)<sup>b</sup> | 620 | 147 (23.7%)<sup>a</sup>, 54 (8.70%)<sup>b</sup> |
| Rea et al. (42) | 2010 | USA | RCT | 1941 | 960 | 296 (30.83%)<sup>a</sup> | 981 | 335 (34.1%)<sup>a</sup> |
| Hallstrom et al. (43) | 2000 | USA | RCT | 520 | 279 | 95 (34.05%)<sup>a</sup> | 241 | 97 (40.2%)<sup>a</sup> |
| Jost et al. (44) | 2011 | France | RCT | 156 | 71 | - | 85 | - |
| Wnent et al. (27) | 2021 | Germany | Prospective cohort | 5406 | 1362 | 174 (12.77%) | 4044 | 320 (7.91%) |
| Schmicker et al. (45) | 2021 | USA | Retrospective cohort | 26810 | 10942 | - | 15868 | - |
| Bohm et al. (46) | 2007 | Sweden | Retrospective cohort | 9354 | 8209 | 591 (0.07%) | 1145 | 77 (6.72%) |
| Iwami et al. (47) | 2007 | Japan | Retrospective cohort | 1327 | 783 | 60 (7.66%) | 544 | 37 (6.80%) |
| Riva et al. (48) | 2019 | Sweden | Observational cohort | 18259 | 11920 | 1647 (13.81%) | 6339 | 853 (13.45%) |
| SOS-KANTO study group (49) | 2007 | Japan | Prospective cohort | 1151 | 712 | - | 439 | - |
| Waalewijn et al. (50) | 2001 | Netherlands | Prospective cohort | 478 | 437 | - | 41 | - |
| Ong et al. (51) | 2008 | Singapore | Prospective cohort | 441 | 287 | 8 (2.78%) | 154 | 4 (2.59%) |
| Bobrow et al. (52) | 2010 | USA | Prospective cohort | 4415 | 666 | - | 849 | - |
| Kitamura et al. (53) | 2018 | Japan | Prospective cohort | 143500 | 41013 | 4,470 (10.89%) | 102487 | 9,899 (9.65%) |
| Javaudin et al. (54) | 2020 | France | Retrospective cohort | 8541 | 1544 | 154 (9.97%) | 6997 | 706 (10.1%) |
| Kaneto et al. (55) | 2024 | Japan | Observational cohort | 7404 | 844 | 46 (5.45%) | 6560 | 109 (1.66%) |
| Olasveengen et al. (56) | 2008 | Norway | Retrospective cohort | 426 | 281 | - | 145 | - |

### Study Selection

A total of 11,186 records were identified through systematic searches in PubMed (n = 2,378), Scopus (n = 1,784), Web of Science (n = 1,300), Embase (n = 2,210), Google Scholar (n = 3,471), and Cochrane Library (n = 43). After removing 981 irrelevant records and 4,142 duplicates, a total of 6,063 studies remained for screening.

During the title and abstract screening, 5,895 records were excluded based on predefined exclusion criteria, including review articles, case reports, non-English articles, non-human studies, and unavailable abstracts. The remaining 168 full-text articles were assessed for eligibility.

After full-text evaluation, 150 studies were excluded due to insufficient data (n = 46), small sample size (n = 7), review/meta-analysis studies (n = 68), and low-quality assessment scores (n = 29). Finally, 18 studies met the inclusion criteria and were incorporated into the meta-analysis, comprising 5 randomized controlled trials (RCTs) (40–44, 57)and 13 observational cohort studies (27, 45–56).

### Characteristics of Included Studies

The 18 selected studies were conducted across various countries, including Sweden, the United States, Japan, Germany, France, and the Netherlands, with a combined sample size of 232,655 OHCA cases. The studies included both RCTs and observational cohorts, with varying sample sizes ranging from 156 to 143,500 participants.

Among the 5 RCTs, three were conducted in Sweden (40, 41, 43), one in the United States (42), and one in France (44). The 13 observational cohort studies were performed in multiple countries (27, 45–56).

### Demographic characteristics

The demographic characteristics of the study populations are described in terms of sample size, gender, age, place of arrest, EMS response time, and epinephrine administration.

The observational and RCT sample sizes are reported, showing a large proportion of male participants in both groups (observational: 49,022 males, RCT: 1,697 males in CPR, 1,633 males in CCO). Female participants were also included in the study, with 31,085 females in the observational group and 879 in the RCT CPR group, 849 in the CCO group. The age range for participants varied between the observational and RCT groups, with the average age being 65.52 years in the observational group and 66.99 years in the RCT group.

### EMS Response Time

The mean time for Emergency Medical Services (EMS) arrival was 7.09 minutes (SD = 4.24) for CPR and 7.44 minutes (SD = 4.47) for CCO, with an overall weighted mean of 7.28 minutes. The difference between the two groups was not statistically significant, indicating a comparable EMS response time across interventions.

### Initial Cardiac Rhythm

The distribution of initial cardiac rhythms varied, with ventricular tachycardia/ventricular fibrillation (VT/VF) being the most common rhythm observed. In the observational group, 11,602 participants presented with VT/VF, while in the RCT group, 813 were in the CPR group and 790 in the CCO group. Other cardiac rhythms included pulseless electrical activity (PEA) and asystole, with PEA observed in 18,323 participants in the observational cohort, and in the RCT group, 99 participants in CPR and 85 in CCO. Asystole was the presenting rhythm in 22,264 observational cases, with 429 in the RCT CPR group and 409 in the CCO group.

### Administration of Epinephrine

Epinephrine administration was reported in 15.45% (n = 6,624) of the CPR group and 19.45% (n = 21,330) of the CCO group, with a total of 18.33% (n = 27,954) across both groups. The slightly higher administration in the CCO group may be attributed to differences in resuscitation response and intervention strategies.

**Table 2.**
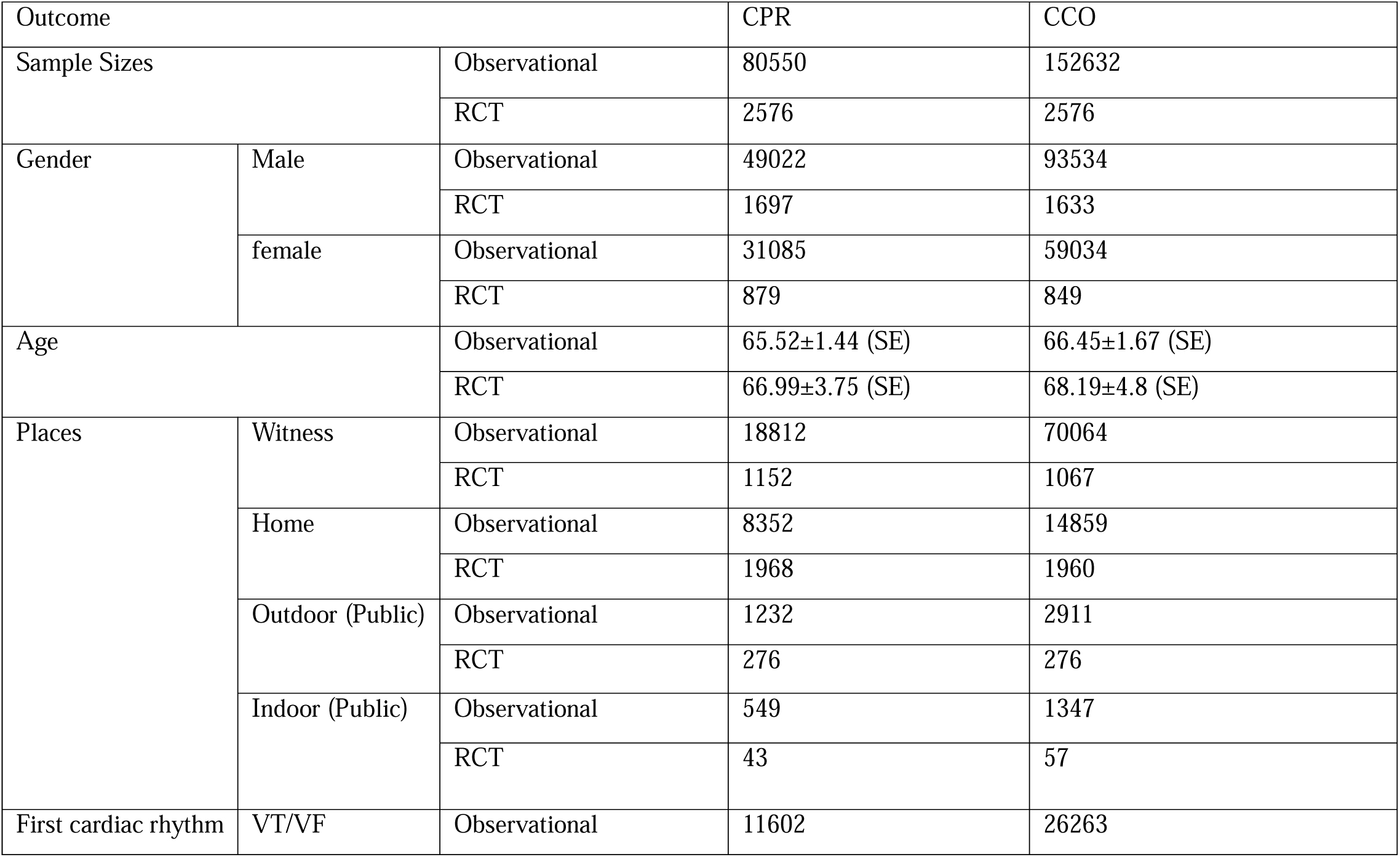
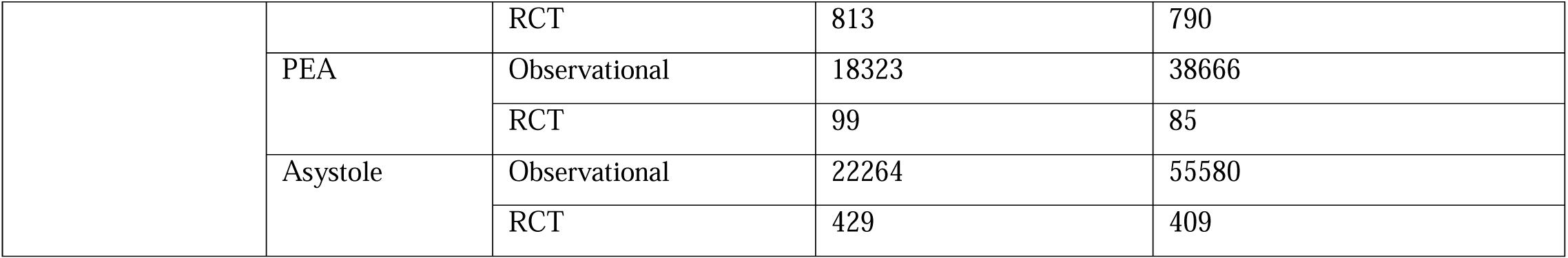
table presents the demographic characteristics of participants in the included studies, categorized by chest compression-only CCO and standard CPR (sCPR). Variables include sample size, gender distribution, mean age, place of arrest, witnessed status, and initial cardiac rhythm, distinguishing between observational and randomized controlled trial (RCT) cohorts. Abbreviations: CCO-CPR, chest compression-only cardiopulmonary resuscitation; sCPR, standard cardiopulmonary resuscitation (compression + ventilation); OHCA, out-of-hospital cardiac arrest; RCT, randomized controlled trial; VT/VF, ventricular tachycardia/ventricular fibrillation; PEA, pulseless electrical activity.

### Prehospital ROSC

There was no significant difference in prehospital ROSC between patients undergoing CPR and those receiving CCO (OR = 1.06; 95% CI: 0.89, 1.27, P = 0.43). The 95% prediction interval was [0.65, 1.73] (Figure 3.A). Heterogeneity across the included studies was high, with an I² value of 82.91%. Sensitivity analysis indicated no significant effect of each study on the overall results (Figure 3.B). The Galbraith plot confirmed high heterogeneity, with Rea, 2010{Rea Thomas, #54} and Wenett, 2021{Wnent, 2021 #95} appearing as outliers (Figure 3.C).

**Figure 3.**
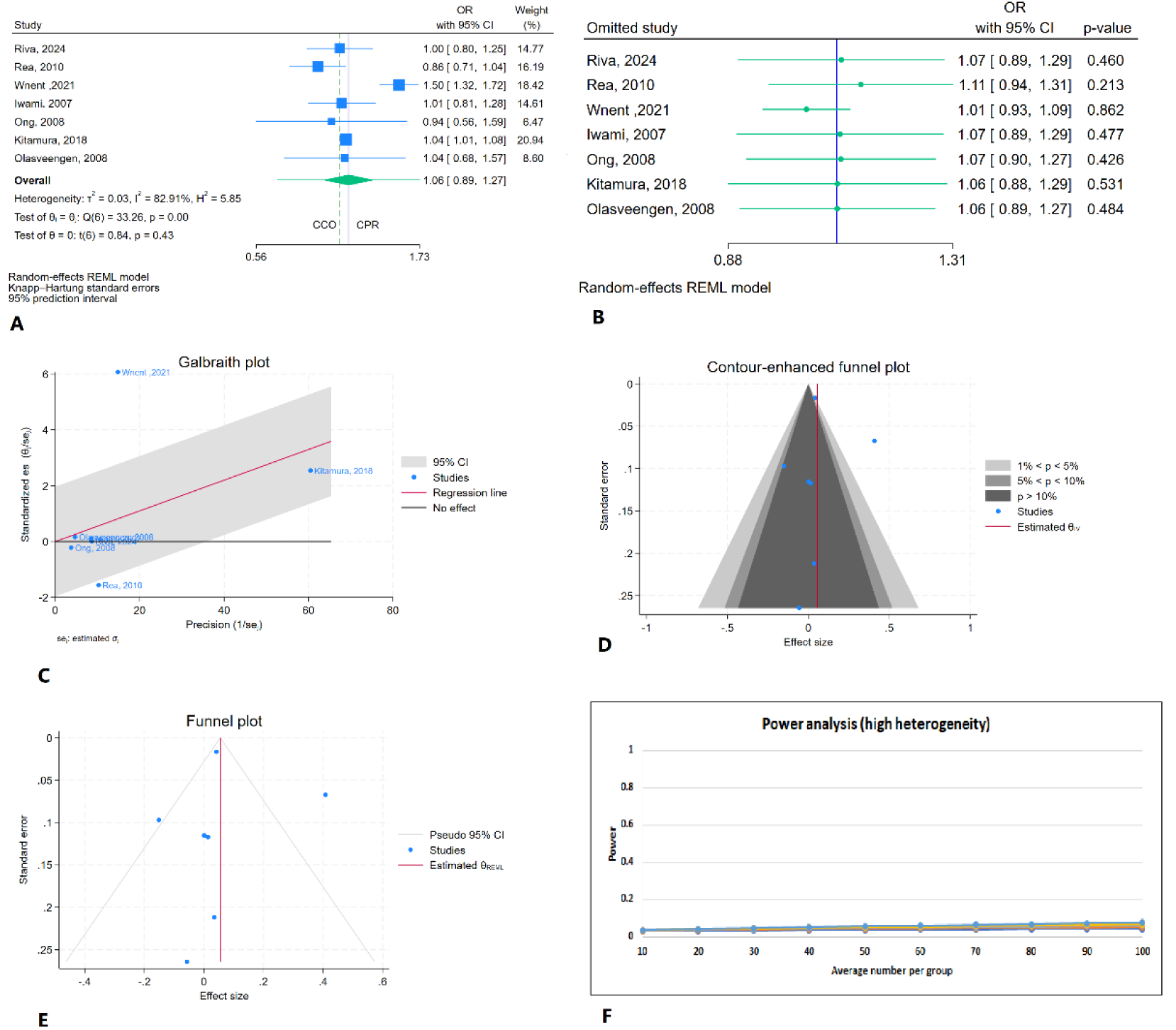
Comparison of prehospital ROSC between CPR and CCO A: Forest plot B: Sensitivity analysis C: Galbraith plot D: Cumulative enhancement funnel plot E: Tin and fill analysis F: Power analysis

Regarding publication bias, the counter-enhanced funnel plot showed low probability of publication bias, which was confirmed by Egger’s test (P = 0.54) and Begg’s test (P = 0.36) (Figure 4.D). The trim-and-fill analysis did not impute any studies (Figure 3.E). The power analysis revealed high statistical power (1 - β = 0.98) (Figure 3.F).

**Figure 4.**
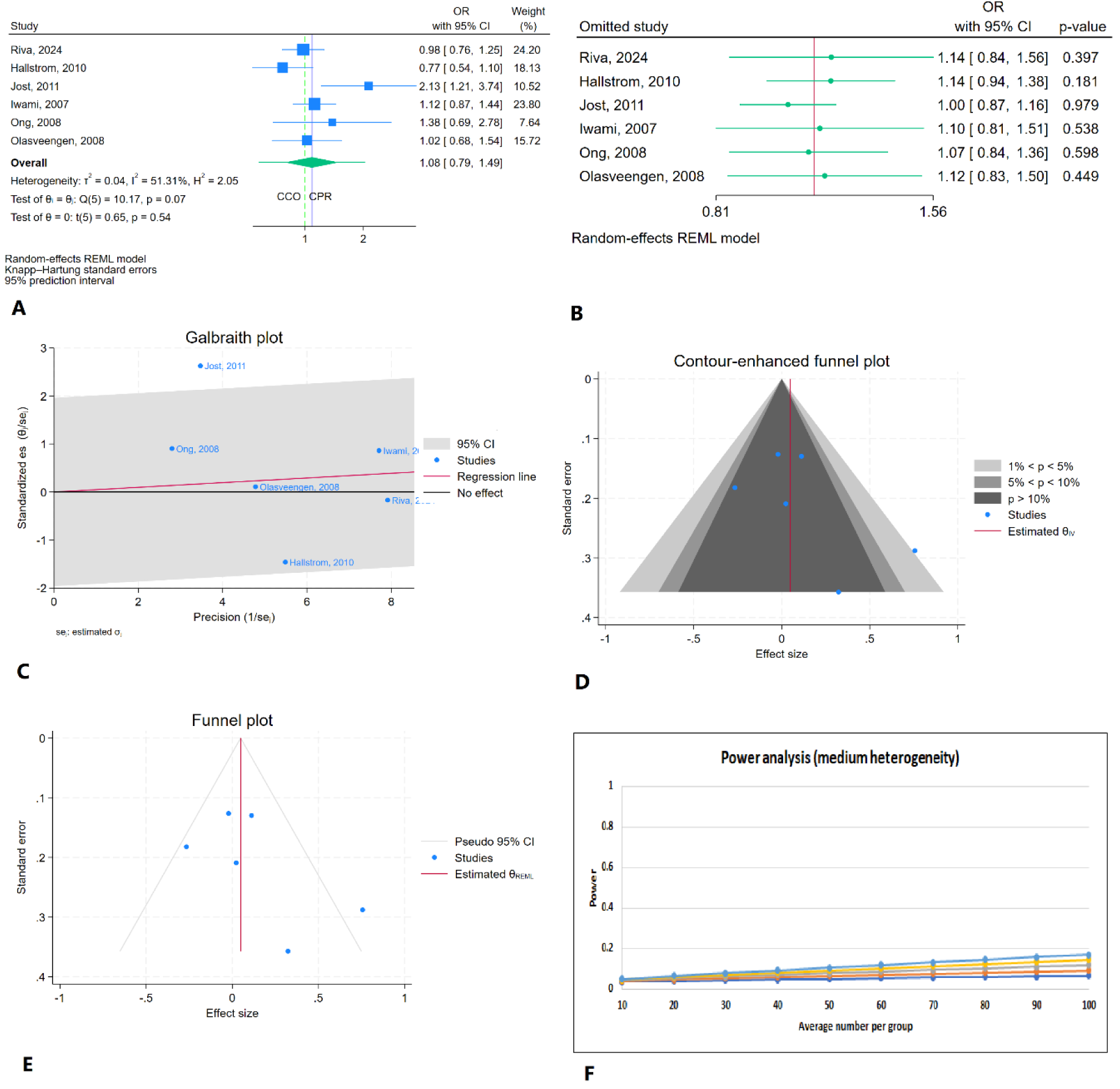
Comparison of in-hospital admission between CPR and CCO A: Forest plot B: Sensitivity analysis C: Galbraith plot D: Cumulative enhancement funnel plot E: Tin and fill analysis F: Power analysis

### In-hospital admission

There was no significant difference in hospital admission between patients undergoing CPR and those receiving CCO (OR = 1.08; 95% CI: 0.79, 1.49, P = 0.54). The 95% prediction interval was [0.59, 1.99] (Figure 4.A). Sensitivity analysis showed no major impact from any single study (Figure 4.B). Heterogeneity across the included studies was moderate, with an I² value of 51.3% (P = 0.07). The Galbraith plot indicated Jost, 2011 as outliers (Figure 4.C)

Regarding publication bias, the contour-enhanced funnel plot showed no indication of asymmetry, which was confirmed by Egger’s test (P = 0.13) and Begg’s test (P = 0.25) (Figure 4.D). The trim-and-fill analysis did not impute any study, indicating no substantial asymmetry in the funnel plot (Figure 4.E). The power analysis revealed low statistical power (1 - β = 0.24), highlighting the need for larger sample sizes (Figure 4.F).

### Survival to Hospital admission after(with) ROSC

There was no significant difference in survival to hospital admission after (with) ROSC between patients undergoing CPR and those receiving CCO (OR = 1.12; 95% CI: 0.53, 2.29, P = 0.34). The 95% prediction interval was [0.01, 118.99] (Figure 5.A). Heterogeneity across the studies included was 89.20%. Sensitivity analysis showed higher survival to hospital admission after (with) ROSC for CPR after the removal of Javaudin, 2020{Javaudin, 2021 #110} (OR = 1.44; 95% CI: 1.13, 1.85, P < 0.01) (Figure 5.B). The Galbraith plot confirmed high heterogeneity, with Javaudin, 2020 {Javaudin, 2021 #110} and Wnent, 2021 {Wnent, 2021 #95} appearing as outliers (Figure 5.C).

**Figure 5.**
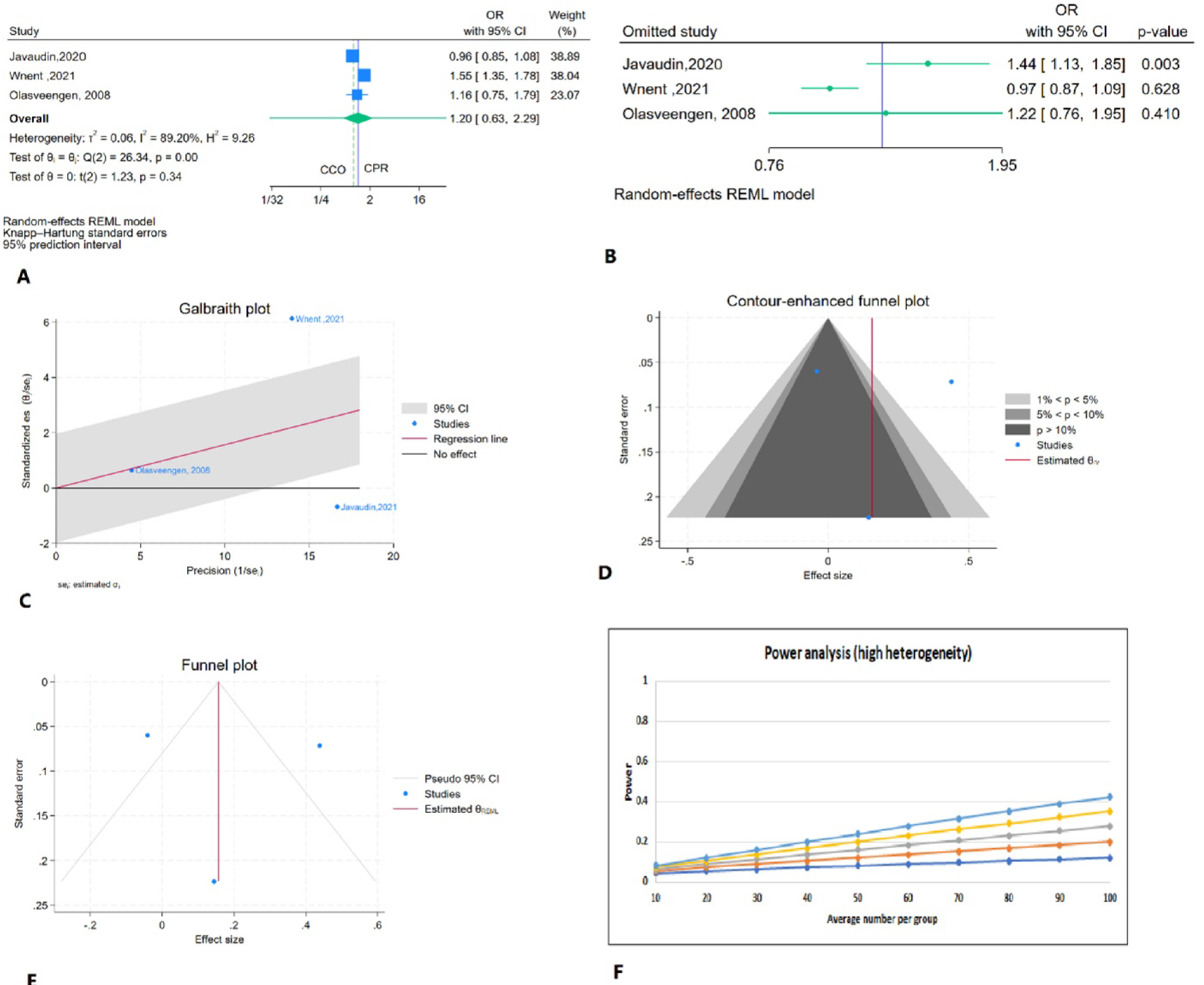
Comparison of Survival to Hospital admission after(with) ROSC between CPR and CCO A: Forest plot B: Sensitivity analysis C: Galbraith plot D: Cumulative enhancement funnel plot E: Tin and fill analysis F: Power analysis

Regarding publication bias, the counter-enhanced funnel plot showed no indication of asymmetry, which was confirmed by Egger’s test (P = 0.96) and Begg’s test (P = 1) (Figure 5.D). The trim-and-fill analysis did not impute any studies (Figure 5.E). The power analysis revealed high statistical power (1 - β = 0.98) (Figure 5.F).

### SHD

There was no significant difference in SHD between patients undergoing CPR and those receiving CCO (OR = 0.85; 95% CI: 0.61, 1.19, P = 0.29). The 95% prediction interval was [0.43, 1.66] (Figure 6.A). Heterogeneity across the included studies was moderate, with an I² value of 66.87%. Sensitivity analysis showed lower SHD in the CPR group after removal of Jost, 2011{Jost, 2011 #100} (OR = 0.79; 95% CI: 0.64, 0.98, P = 0.03) (Figure 6.B). The Galbraith plot confirmed moderate heterogeneity, with Jost, 2011{Jost, 2011 #100} as an outlier (Figure 6.C).

**Figure 6.**
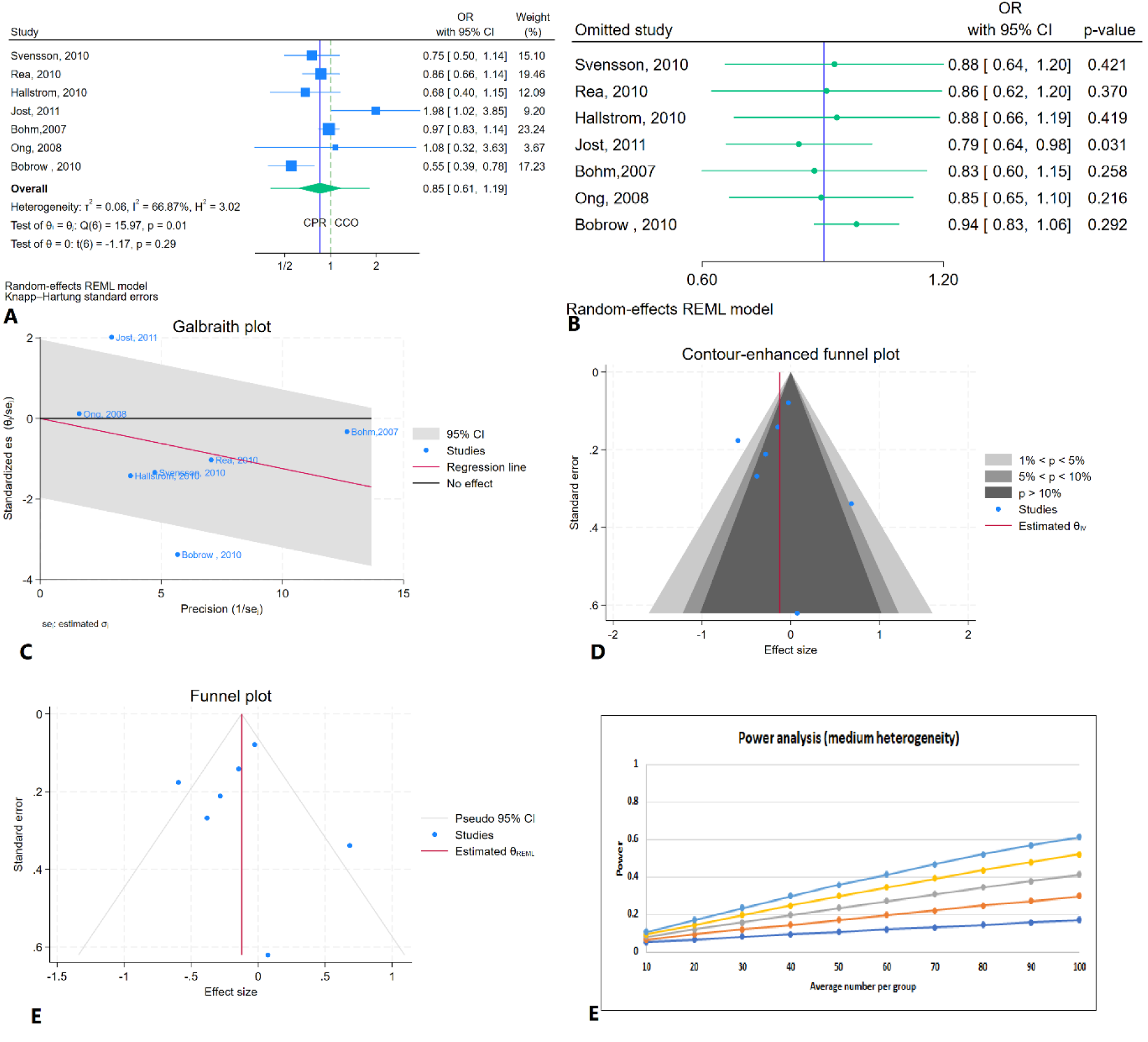
Comparison of SHD between CPR and CCO A: Forest plot B: Sensitivity analysis C: Galbraith plot D: Cumulative enhancement funnel plot E: Tin and fill analysis F: Power analysis

**Figure 7.**
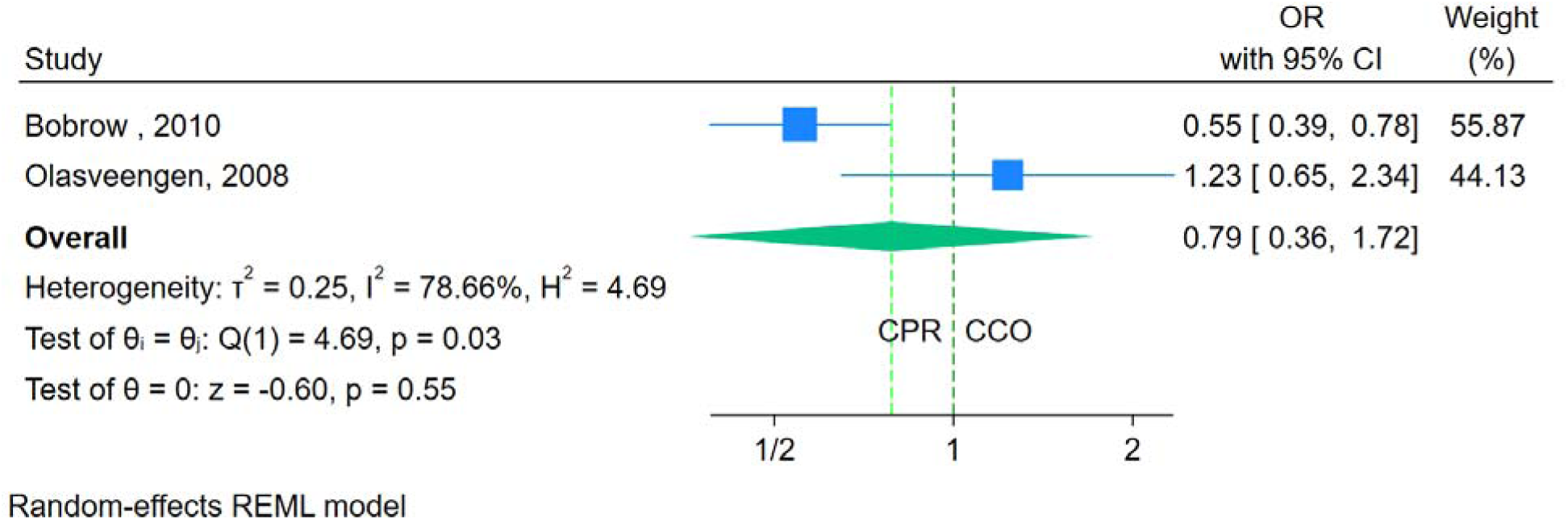
Comparison of hospital discharge between CPR and CCO

Regarding publication bias, the counter-enhanced funnel plot showed no presence of publication bias, which was confirmed by Egger’s test (P = 0.48) and Begg’s test (P = 1) (Figure 6.D). The trim-and-fill analysis did not impute any studies (Figure 6.E). The power analysis revealed high power (1 - β = 0.99) (Figure 6.F).

### Hospital discharge

There was no significant difference in hospital discharge between patients undergoing CPR and those receiving CCO (OR = 0.79; 95% CI: 0.36, 1.72, P = 0.55). However, there were only two studies for this point, so further assessment was not possible.

### SHD with favorable neurological outcome

There was no significant difference in SHD with favorable neurological outcome between patients undergoing CPR and those receiving CCO (OR = 1.47; 95% CI: 0.09, 22.68, P = 0.61) (Figure 8.A). Heterogeneity across the included studies was high, with an I² value of 95.54%. Sensitivity analysis indicated no significant effect of each study on the overall results (Figure 8.B). The Galbraith plot identified Kaneto, 2024{Kaneto, 2024 #111}, and Rea 2010 {Rea Thomas, 54}as outliers (Figure 8.C).

**Figure 8.**
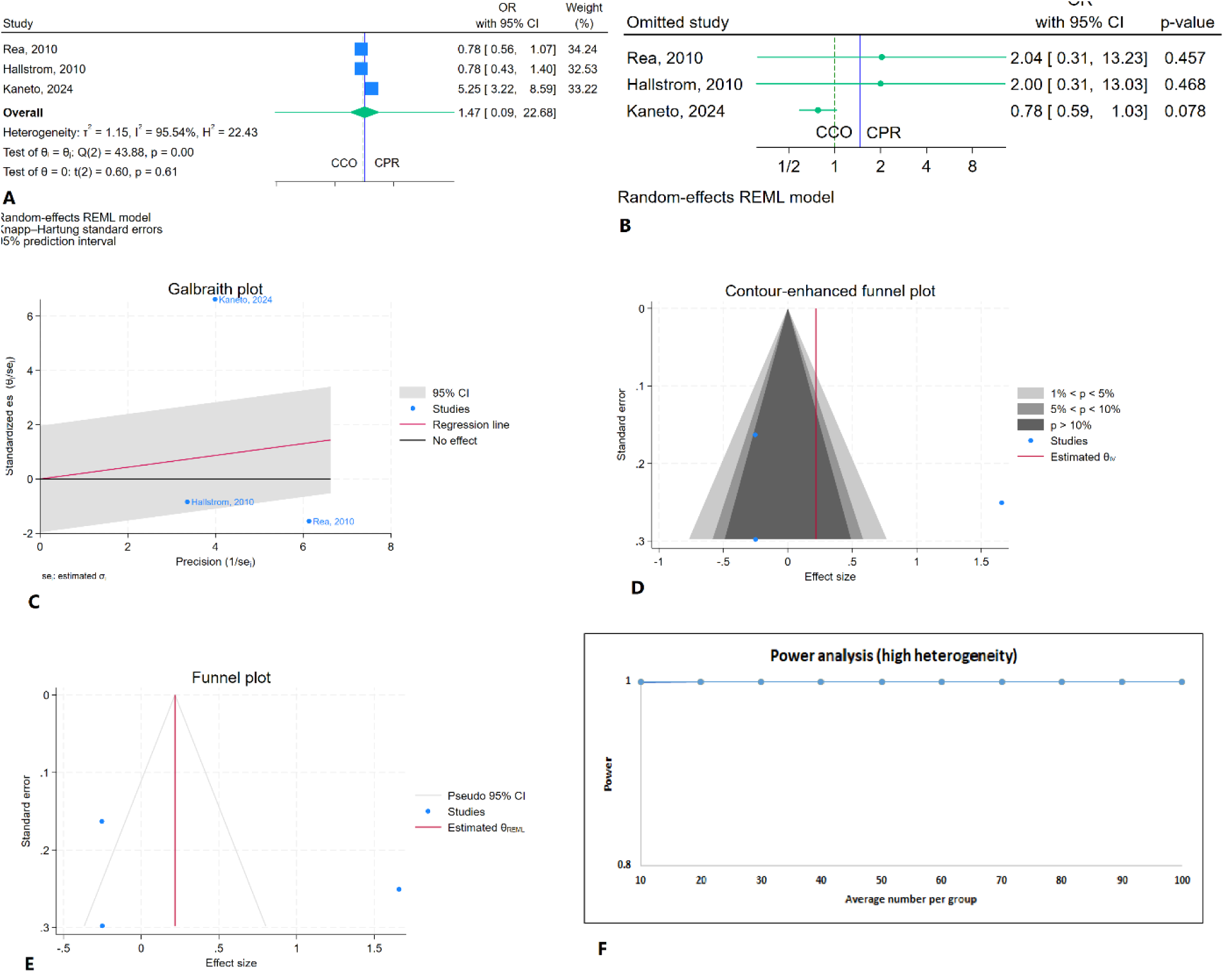
Comparison of SHD with favorable neurological between CPR and CCO

Regarding publication bias, the funnel plot showed no indication of asymmetry, which was confirmed by Egger’s test (P = 0.85) and Begg’s test (P = 1) (Figure 8.D). The trim-and-fill analysis did not impute any studies (Figure 8.E). The power analysis revealed high statistical power (1 - β = 1) (Figure 8.F).

### Overall favorable neurologic outcome

There was no significant difference in overall favorable neurologic outcome between patients undergoing CPR and those receiving CCO (OR = 0.87; 95% CI: 0.64, 1.20, P = 0.32). The 95% prediction interval was [0.43, 1.58] (Figure 9.A). Heterogeneity across the included studies was low, with an I² value of 34.57%. Sensitivity analysis indicated no significant effect of each study on the overall results (Figure 9.B). The Galbraith plot identified no study as outlier (Figure 9.C).

**Figure 9.**
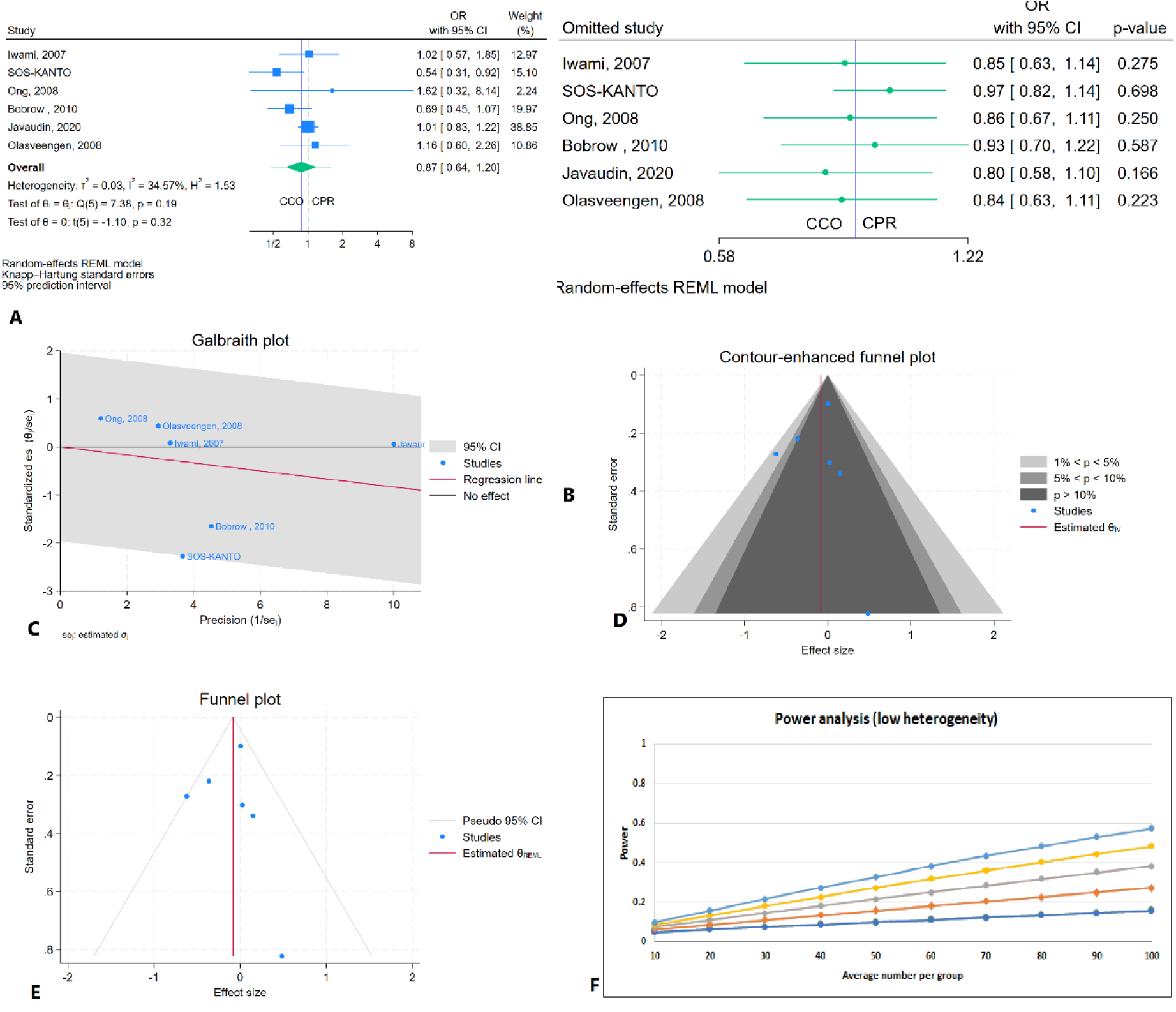
Comparison of Overall favorable neurologic between CPR and CCO

Regarding publication bias, the funnel plot showed no indication of asymmetry, which was confirmed by Egger’s test (P = 0.69) and Begg’s test (P = 1) (Figure 9.D). The trim-and-fill analysis did not impute any studies (Figure 9.E). The power analysis revealed high statistical power (1 - β = 0.99) (Figure 9.F).

### 24-hour mortality

There was no significant difference in 24-hour mortality between patients undergoing CPR and those receiving CCO (OR = 0.92; 95% CI: 0.83, 1.01, P = 0.07). The 95% prediction interval was [0.81, 1.02] (Figure 10.A). Heterogeneity across the included studies was low, with an I² value of 0.00%. Sensitivity analysis showed lower 24-hour survival rates for CPR after the removal of Riva, 2024 {Riva, 2024 #96}(OR = 0.90; 95% CI: 0.82, 1.00, P = 0.04) and Javaudin, 2020 {Javaudin, 2021 #110}(OR = 0.87; 95% CI: 0.77, 0.99, P = 0.02) (Figure 10.B). The Galbraith plot confirmed no outliers (Figure 10.C). Regarding publication bias, the funnel plot showed no indication of asymmetry, which was confirmed by Egger’s test (P = 0.73) and Begg’s test (P = 0.22) (Figure 10.D). The trim-and-fill analysis did not impute any studies (Figure 10.E). The power analysis revealed high statistical power (1 - β = 0.90) (Figure 10.F).

**Figure 10.**
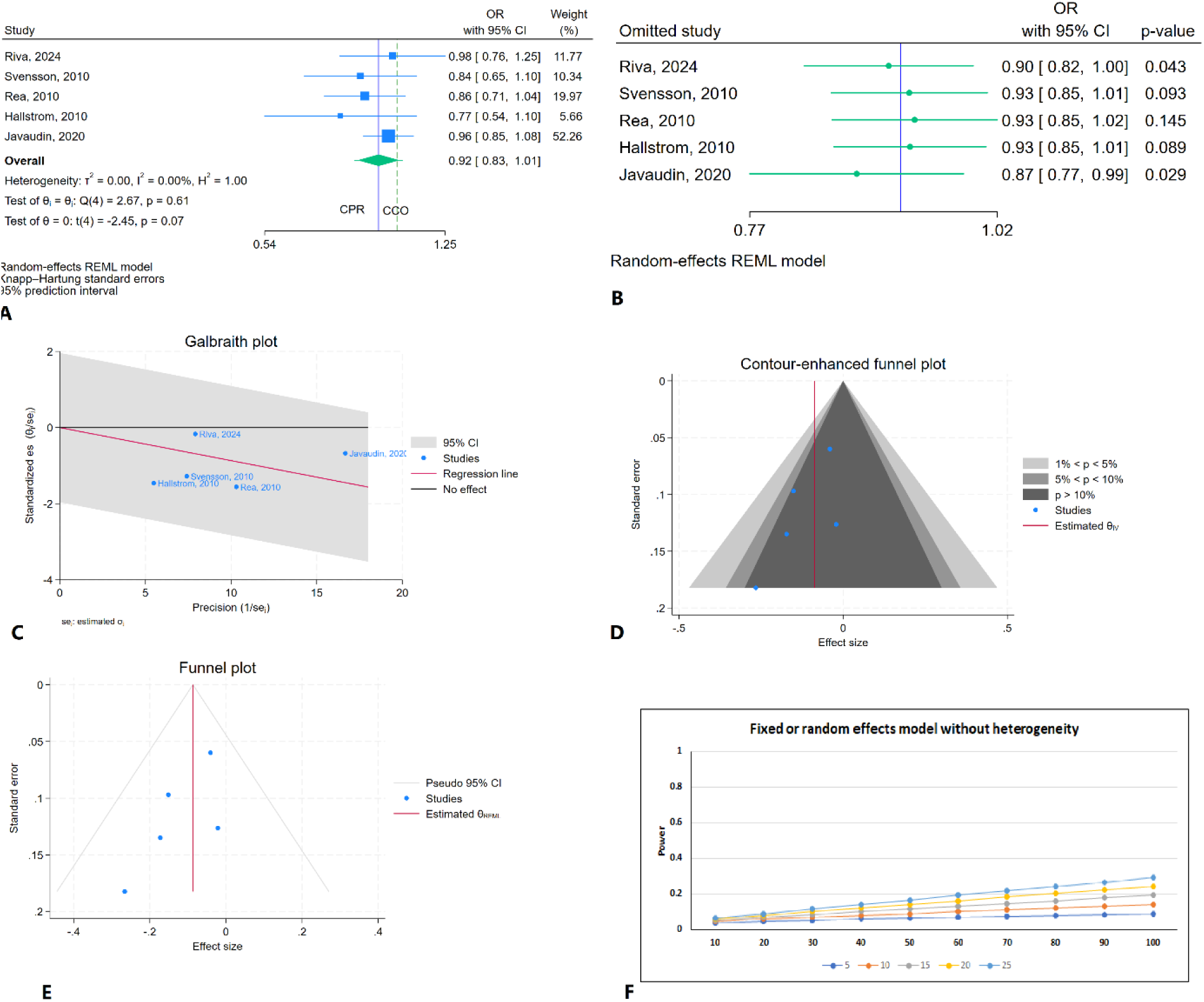
Comparison of 24-hour mortality between CPR and CCO A: Forest plot B: Sensitivity analysis C: Galbraith plot D: Cumulative enhancement funnel plot E: Tin and fill analysis F: Power analysis

### One-Month Mortality

There was no significant difference in one-month mortality between patients undergoing CPR and those receiving CCO (OR = 1.26; 95% CI: 0.98, 1.62, P = 0.07). The 95% prediction interval was [0.51, 3.08] (Figure 11.A). Heterogeneity across the included studies was high, with an I² value of 95.67%%. Sensitivity analysis showed higher one month mortality after removal of Svensson (OR = 1.32; 95% CI: 1.02, 1.71, P = 0.03) (Figure 11.B). The Galbraith plot identified Kaneto, 2024{Kaneto, 2024 #111}, Wenet, 2021{Wnent, 2021 #95}, and Riva 2019{Riva, 2019 #22} as outliers (Figure 11.C).

**Figure 11.**
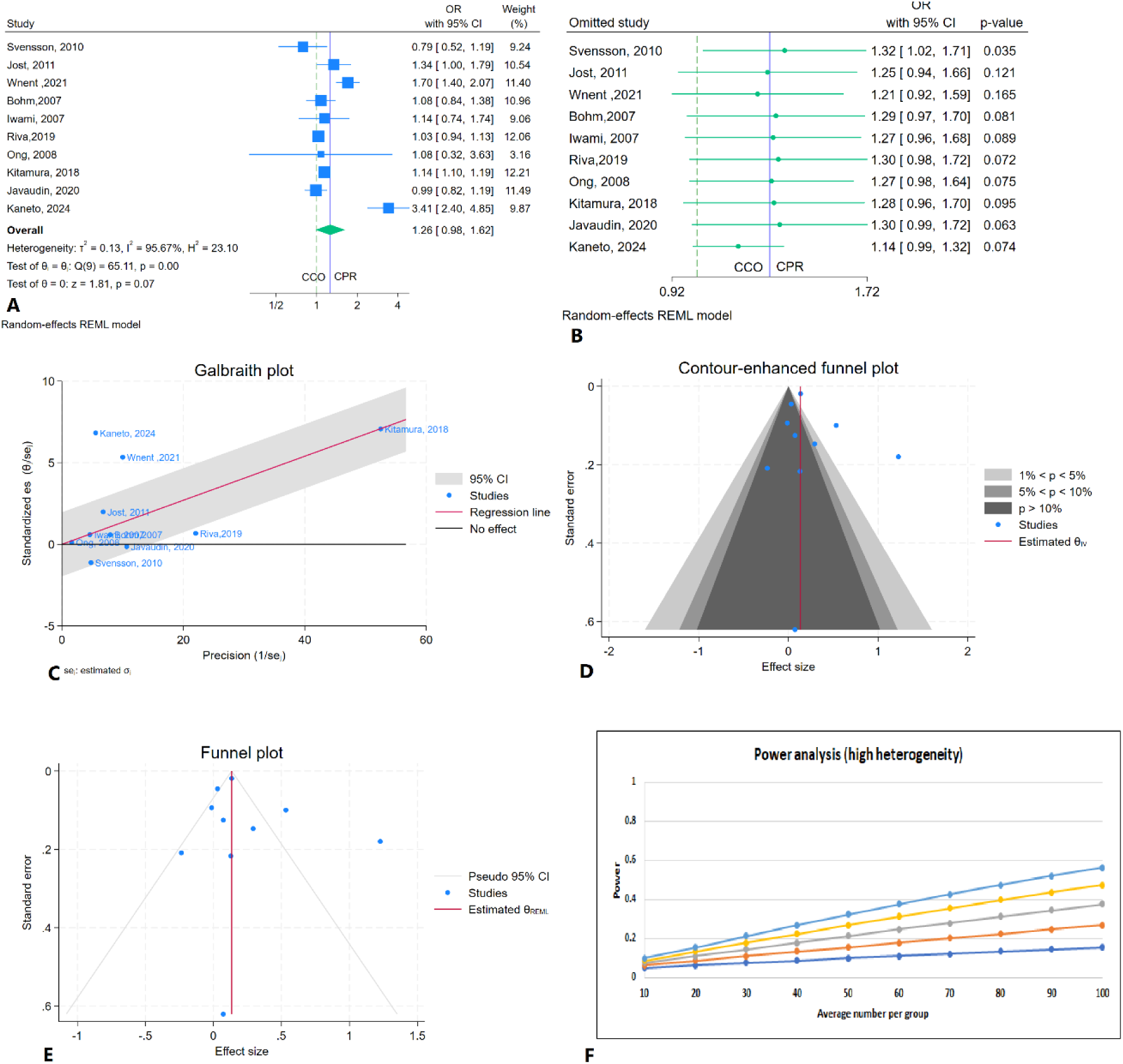
Comparison of One-Month Mortality between CPR and CCO A: Forest plot B: Sensitivity analysis C: Galbraith plot D: Cumulative enhancement funnel plot E: Tin and fill analysis F: Power analysis

Regarding publication bias, the funnel plot showed no indication of asymmetry, which was confirmed by Egger’s test (P = 0.91) and Begg’s test (P = 0.59) (Figure 11.D). The trim-and-fill analysis did not impute any studies (Figure 11.E). The power analysis revealed high statistical power (1 - β = 1) (Figure 11.F).

### Results of the subgroup analysis

The results of the subgroup analysis are presented across various outcomes, sample sizes, and statistical measures. For in-hospital admission, the observational subgroup included 3 studies, showing an odds ratio (OR) of 1.11 (95% CI: 0.90, 1.36) with low heterogeneity (I² = 0.00%). The RCT subgroup, with 3 studies, showed an OR of 1.12 (95% CI: 0.64, 1.95) with higher heterogeneity (I² = 85.04%). For SHD, the observational group showed an OR of 0.78 (95% CI: 0.50, 1.24) with moderate heterogeneity (I² = 75.47%), while the RCT subgroup had an OR of 0.91 (95% CI: 0.63, 1.31) with moderate heterogeneity (I² = 62.60%).

Regarding SHD with a good neurological outcome, the observational group had an OR of 1.36 (95% CI: 0.70, 2.66) with high heterogeneity (I² = 97.33%), while the RCT group showed a lower OR of 0.77 (95% CI: 0.58, 1.02) with no heterogeneity (I² = 0.00%). For prehospital ROSC, the observational group had an OR of 1.13 (95% CI: 0.93, 1.37) with high heterogeneity (I² = 82.88%), while the RCT group showed an OR of 0.92 (95% CI: 0.79, 1.06) with low heterogeneity (I² = 1.99%).

The 24-hour mortality analysis in the observational group showed an OR of 0.96 (95% CI: 0.85, 1.08), with low heterogeneity (I² = 0.00%). In the RCT subgroup, the OR was 0.87 (95% CI: 0.77, 0.98) with no heterogeneity (I² = 0.00%). One-month mortality in the observational group showed an OR of 1.32 (95% CI: 0.98, 1.78) with high heterogeneity (I² = 96.77%), and in the RCT group, the OR was 1.05 (95% CI: 0.62, 1.76) with moderate heterogeneity (I² = 76.57%). These subgroup analyses provide valuable insights into the effects of sCPR versus CCO on various outcomes across different patient subgroups.

**Table 3.**
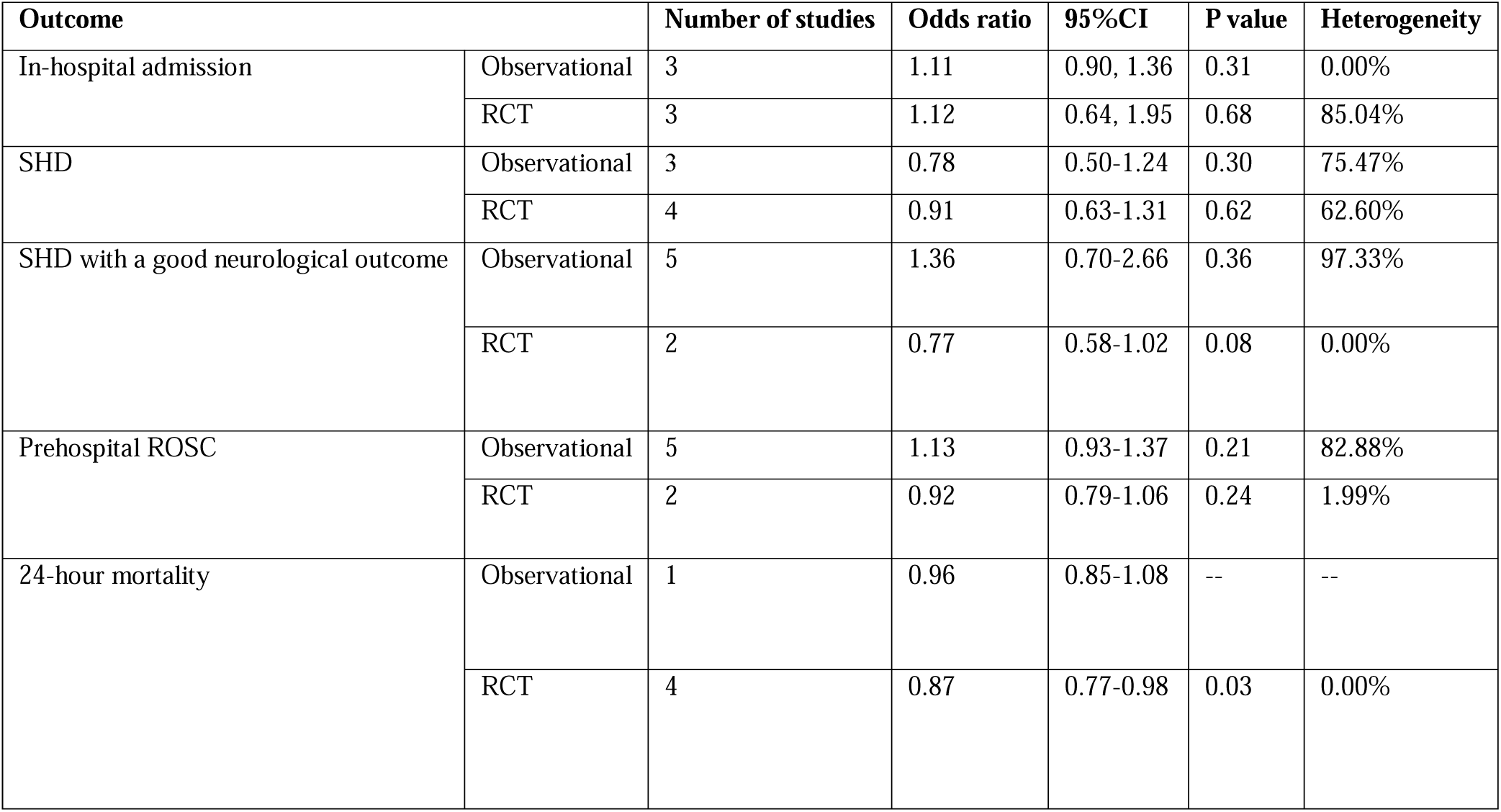
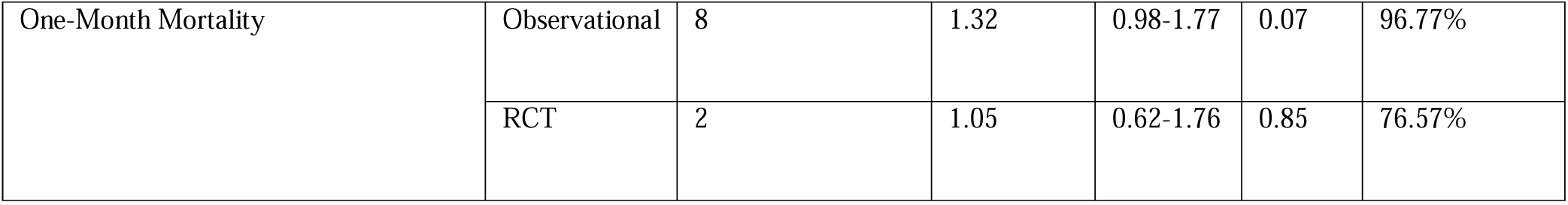
Comparison of the effects of chest compression-only CCO vs. standard CPR (sCPR) in different study designs on survival to hospital discharge (SHD), neurological outcomes, return of spontaneous circulation (ROSC), and mortality outcomes. Results are presented as odds ratios (OR) with 95% confidence intervals (CI), P-values, and heterogeneity estimates, stratified by randomized controlled trials (RCTs) and observational studies. Abbreviations: OHCA – Out-of-Hospital Cardiac Arrest; CPR – Cardiopulmonary Resuscitation; CCO-CPR – Chest Compression-Only Cardiopulmonary Resuscitation; sCPR – Standard Cardiopulmonary Resuscitation (compression + ventilation); SHD – Survival to Hospital Discharge; ROSC – Return of Spontaneous Circulation; RCT – Randomized Controlled Trial; OR – Odds Ratio; CI – Confidence Interval

### Quality Assessment of Studies

The quality assessment showed that all observational studies were high quality, scoring 8–9 on the Newcastle-Ottawa Scale (NOS). RCTs varied in quality, with two having low risk of bias, one with some concerns, and two with high risk due to issues in randomization and outcome reporting. Overall, the observational studies were strong, while some RCTs had limitations that may affect reliability. These findings support confidence in observational data but call for caution when interpreting lower-quality RCTs (*Supplementary Table 4. & 5.*).

## Discussion

Our meta-analysis found no significant differences between CCO and sCPR in key survival and resuscitation outcomes, including in-hospital admission, survival to hospital discharge (SHD), neurological recovery, prehospital ROSC, and survival rates at 24 hours and 1 month. Notably, prior meta-analyses did not examine the 24-hour and 1-month survival periods separately, making our findings more comprehensive(1). Regarding neurological outcomes, both CPR methods yielded similar results, with no significant difference in overall neurological recovery (OR = 0.87, 95% CI: 0.64–1.20, P = 0.32) or SHD with favorable neurological outcomes (OR = 1.47, 95% CI: 0.09–22.68, P = 0.61). For mortality outcomes, 24-hour survival was comparable between CCO and sCPR (OR = 0.92, 95% CI: 0.83–1.01, P = 0.07). One-month mortality also showed no significant difference (OR = 1.26, 95% CI: 0.98–1.62, P = 0.07), but sCPR carried a slightly higher risk after outlier analysis (OR = 1.32, 95% CI: 1.02–1.71, P = 0.03). While neither CPR method demonstrated clear superiority, these findings reinforce CCO as an effective and practical approach for bystander resuscitation, particularly given its ease of execution.

Our study revealed no additional survival benefits associated with mouth-to-mouth or any rescue oxygenation or generally ventilation framing CCO as the most efficient strategy. Bystander-initiated CPR is a time-sensitive intervention that significantly impacts survival in out-of-hospital cardiac arrest (2). Like sCPR, chest compression is more efficient when done by trained professionals, (3) but remains a significant barrier for bystanders (4). Therefore, this method addresses all the challenges posed by logistical as well as practical barriers during the cardiac arrest emergency setting. These challenges make CCO the more practical, and scalable method for bystander sCPR.

One of the toughest barriers in the provision of bystander sCPR is the aversion in performing mouth to mouth ventilation(5), especially on strangers (6). This reluctance stems from concerns such as the risk of contracting diseases (7), physical intimacy which creates discomfort in performing ventilation properly (8), which in turn may result in lack of proper ventilation as well as disturbed chest compression. (9). Consequently, many people who may be inclined to help do not wish to perform sCPR in situations where ventilation is needed (10, 11). According to studies, when dispatchers instruct callers to perform CCO, bystander engagement increases considerably (12, 13). Given that early sCPR initiation is one of the strongest predictors of survival, implementing an approach that motivates more individuals to intervene is critical for improving out-of-hospital cardiac arrest outcomes (14–16).

Another factor that should be considered are the physiological considerations of cardiac arrest. Generally, cardiac arrest of organic origin like cardiac causes, such as ventricular fibrillation or ventricular tachycardia are significant causes of cardiac arrest which could be effectively resuscitated by external shock (17, 18). We found that 21% of total arythmias were shockable, including VF or VT, in comparison to previous study which was overall proportion of a shockable initial rhythm about 38%–37% (19).

During the first few minutes after collapse, the oxygen supply within the bloodstream is sufficient (20, 21), so maintaining an uninterrupted circulatory effort is the primary objective (22, 23). Vital organ perfusion can be maintained during chest compressions (24, 25). However, intermission for mouth-to-mouth results in cessation of chest compressions leading to reduced blood flow, coronary and cerebral perfusion pressure, and spontaneous circulation. In this regard, several studies have shown even lower survival rates in interrupted as compared to sustained chest compression (26–28). In our meta-analysis, we found that sCPR techniques and CCO method contributed to similar prehospital ROSC rates. That is, CCO is sufficient for adequate oxygen supply in the critical early minutes of resuscitation, considering the time interval between EMS arrival at the cardiac arrest site. The prehospital time frame is crucial, as the average time interval between EMS arrival at the cardiac arrest site was 7.28 minutes. From these findings, CCO might be recommended for pre-arrival of emergency medical services.

Bystanders who lack sufficient medical training face the additional challenge of integrating the performing compressions and ventilations (3, 29, 30). This is could be challenge particularly acute for dispatcher-assisted CPR situations, where a dispatcher instructs a bystander in real-time (31). Teaching CCO allows for quicker onset of compression, minimization of cognitive burden, and greater adherence to dispatcher’s instructions, as it is easier to understand and follow (32–34). This leads to a higher tendency of effective rescuer performance. On the other hand, asking a layman to provide ventilation opens up several other steps like mouth sealing, airway opening, and managing hyperventilation (35). Such issues can result in obstructed or delayed CPR and thus worsen the chance of survival (35). Even when bystanders provide rescue breaths, their attempts are often ineffective due to inadequate airway positioning/hyperventilation, ineffective sealing of the airway, and increased gastric inflation, which leads to aspiration (9, 29, 35). Moreover, public training campaigns can better promote the performance of CCO as they are more likely to be utilized by untrained people (36–38).

The interval between the onset of the cardiac arrest, arrival of the EMS, and the provision of CPR and resuscitation differs depending on the area, traffic conditions, and the effectiveness of Emergency Medical Services resources (39). As we found, the mean time of EMS arrival is 7.28 minutes. While urban EMS has response times of less than eight minutes, rural areas are often overwhelmed with delays (41, 40). During this gap of time, circulation maintenance remains the most vital factor to be focused on in increasing the chances of survival. Bystanders often administer CPR for only a few minutes before EMS arrives; therefore, prioritizing continuous chest compressions provides maximum perfusion during this vital window (22, 42). Ventilation, on the other hand, becomes critical only in extended resuscitations, which are often administered by experienced EMS staff rather by bystanders (43).

Our findings are consistent with previous clinical trials and meta-analyses, which have found no substantial superiority of sCPR over CCO regarding survival rate. Some studies demonstrated that CCO is even associated with better outcomes, especially in cardiac-origin OHCA. For example, Hallstrom et al. reported that CCO was associated with 29% increased survival in comparison to sCPR, though it was not statistically significant (44). Rea et al. discovered that patients with shockable rhythms received greater benefit from CCO than sCPR (45). Furthermore, Swenson et al. found that bystander-initiated ventilation did not improve survival (45). Given that our study included a bigger sample size and a larger dataset than prior meta-analyses (1), our findings improve the evidence for CCO as the preferred bystander CPR approach.

The CCO group has 152,632 patients, the sCPR group has 80,550, and the RCT groups have 2,576 each. This collection of data will enable more rigorous stratified findings to gain additional insights into CPR practices and strategies, even larger data, and a more investigated study than the previous meta-analysis (1). The condition of observing all possible related demographic factors, such as any of whom could have a variety of cardiac activity (such as VT/VF, PEA, or asystole) in any number of contexts, makes the study applicable to real-world OHCA scenarios. Furthermore, unlike previous studies, our study examines two periods of survival outcomes, such as 24 hours and one month survival, providing more options for evaluating the effectiveness of CPR in participants, and also we analyzed the neurological outcome separately, including during discharge status and neurological examination of patients in studies that had follow-up, which were not observed in prior similar meta-analysis (1).

To ensure statistical robustness, we used heterogeneity assessment, sensitivity analysis, and publication bias testing (trim-and-fill analysis, Egger’s and Begg’s tests, and cumulative funnel plots), which confirmed the stability and absence of bias. We also performed a sensitivity analysis, which found no disproportionate impact of any singular study. Collectively, these pieces of evidence further support the credibility of our conclusions. This meta-analysis provides actionable data for strategies for dispatcher-assisted CPR techniques and training programs for bystanders. In particular, it promotes CCO to be the leading intervention to encourage bystanders.

As the willingness to perform CCO is much higher in the population, and it is much easier to perform and teach, CCO is an ideal resuscitation method that should be strongly advocated in the guidelines, which can lead to numerous lifesavings.

### Limitations

This meta-analysis has several limitations that should be considered. The limited number of RCTs means that findings rely heavily on observational data, which is more prone to confounding and selection bias, despite performing sensitivity analyses, heterogeneity assessments, and bias detection tests, the potential for residual confounding cannot be eliminated. Additionally, methodological variability across studies, including differences in CPR quality, bystander training, dispatcher-assisted CPR, and EMS response times, may have influenced outcomes.

Variability in EMS response times is a key factor, as CCO showed potential benefits with an average EMS arrival time of 7.28 minutes, but response times differ by geography and healthcare efficiency, affecting real-world applicability. Lack of standardized outcome reporting, particularly for long-term neurological recovery, also limits conclusions, as many studies did not follow patients beyond hospital discharge.

Publication and selection biases remain concerns, though trim-and-fill analysis, Egger’s test, and Begg’s test showed no significant bias. The differences in study population characteristics, such as age, comorbidities, and initial cardiac rhythm, may also impact generalizability. Moreover, differences in public health infrastructure, AED availability, and EMS protocols across regions could have affected results.

Variability in bystander CPR training rates, EMS protocols, and access to early defibrillation could impact the relative effectiveness of CCO versus sCPR in real-world settings. Despite these limitations, our large sample size, rigorous statistical methodology, and inclusion of diverse OHCA scenarios provide a robust evidence base for evaluating the effectiveness of CCO compared to sCPR. Future research should prioritize large-scale, high-quality RCTs with longer follow-up durations to assess neurological and functional recovery beyond hospital discharge. Additionally, further subgroup analyses examining EMS response times, arrest etiologies, and bystander training levels may help refine our understanding of when CCO may be most effective.

## Conclusion

Despite no statistically significant survival difference between CCO and sCPR, multiple practical, physiological, and public health considerations, CCO might be superior approach for bystanders. It increases bystander willingness, ensuring higher CPR initiation rates, providing uninterrupted perfusion to the brain and heart, is easier to teach, learn, and perform, both in public training and dispatcher-assisted CPR. Ventilation by laypersons is frequently ineffective or harmful, making it non-essential in most cases. Faster response times and simpler execution make CCO the most feasible and practical method. Given our larger sample size, broader analysis, and strong statistical foundation, our study provides robust evidence for modifying CPR guidelines. Future research should continue to explore long-term survival benefits, while public health initiatives should prioritize training more bystanders in CCO to improve out-of-hospital cardiac arrest survival rates worldwide.

## Data Availability

Data is Publicly available as meta Analysis

## Abbreviation list

OHCA: Out-of-Hospital Cardiac Arrest
CPR: Cardiopulmonary Resuscitation
CCO-CPR: Chest Compression-Only Cardiopulmonary Resuscitation
sCPR: Standard Cardiopulmonary Resuscitation (compression + ventilation)
SHD: Survival to Hospital Discharge
ROSC: Return of Spontaneous Circulation
RCT: Randomized Controlled Trial
OR: Odds Ratio
CI: Confidence Interval
EMS: Emergency Medical Services
AED: Automated External Defibrillator
VF: Ventricular Fibrillation
VT: Ventricular Tachycardia
PEA: Pulseless Electrical Activity
NOS: Newcastle-Ottawa Scale
ROB-2: Revised Cochrane Risk-of-Bias Tool
BLS: Basic Life Support
AHA: American Heart Association
ERC: European Resuscitation Council
CPC: Cerebral Performance Category

## Founding

None

## Competing interests

None

## Supplementary Materials

**Supplementary Table 1.**
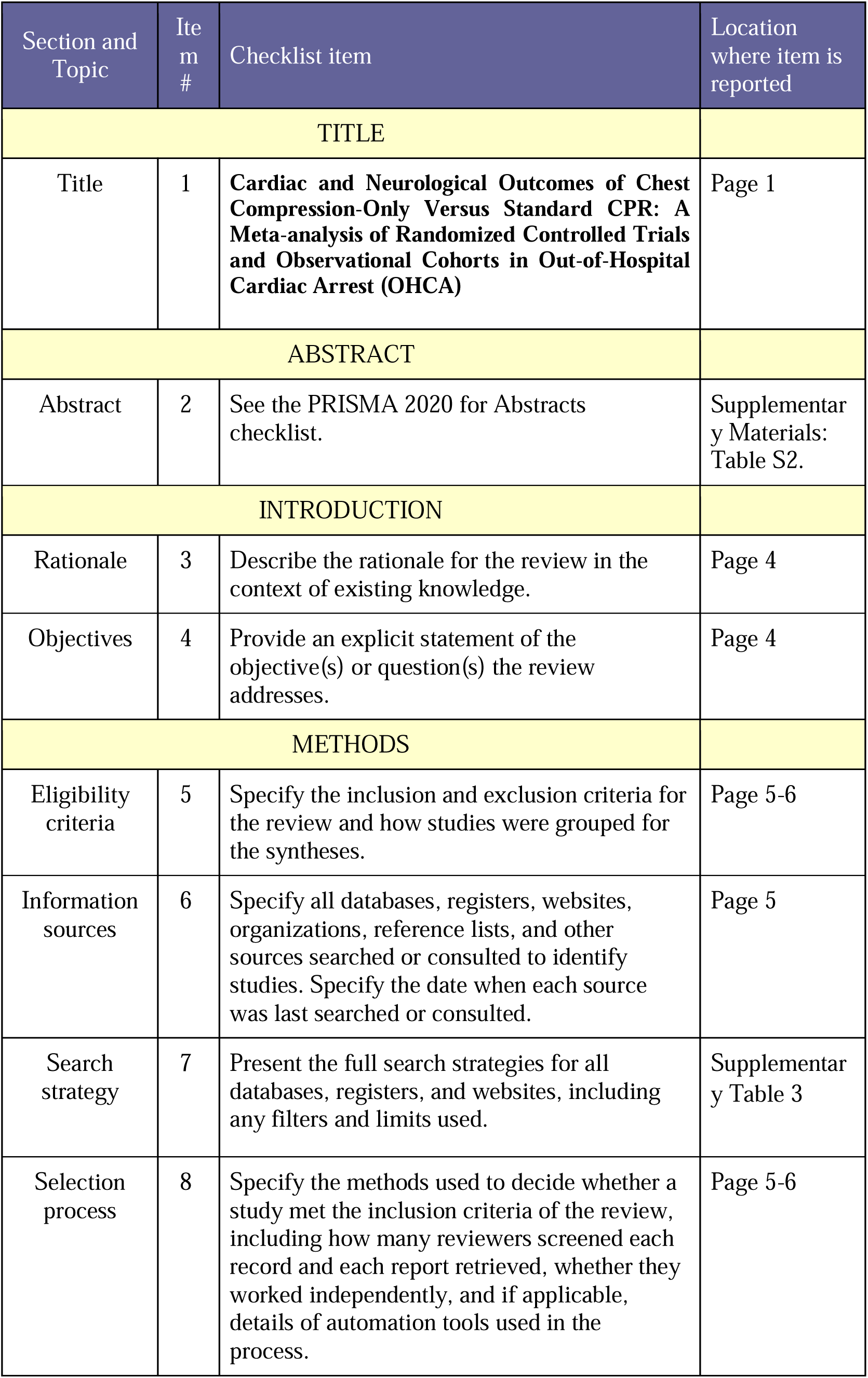
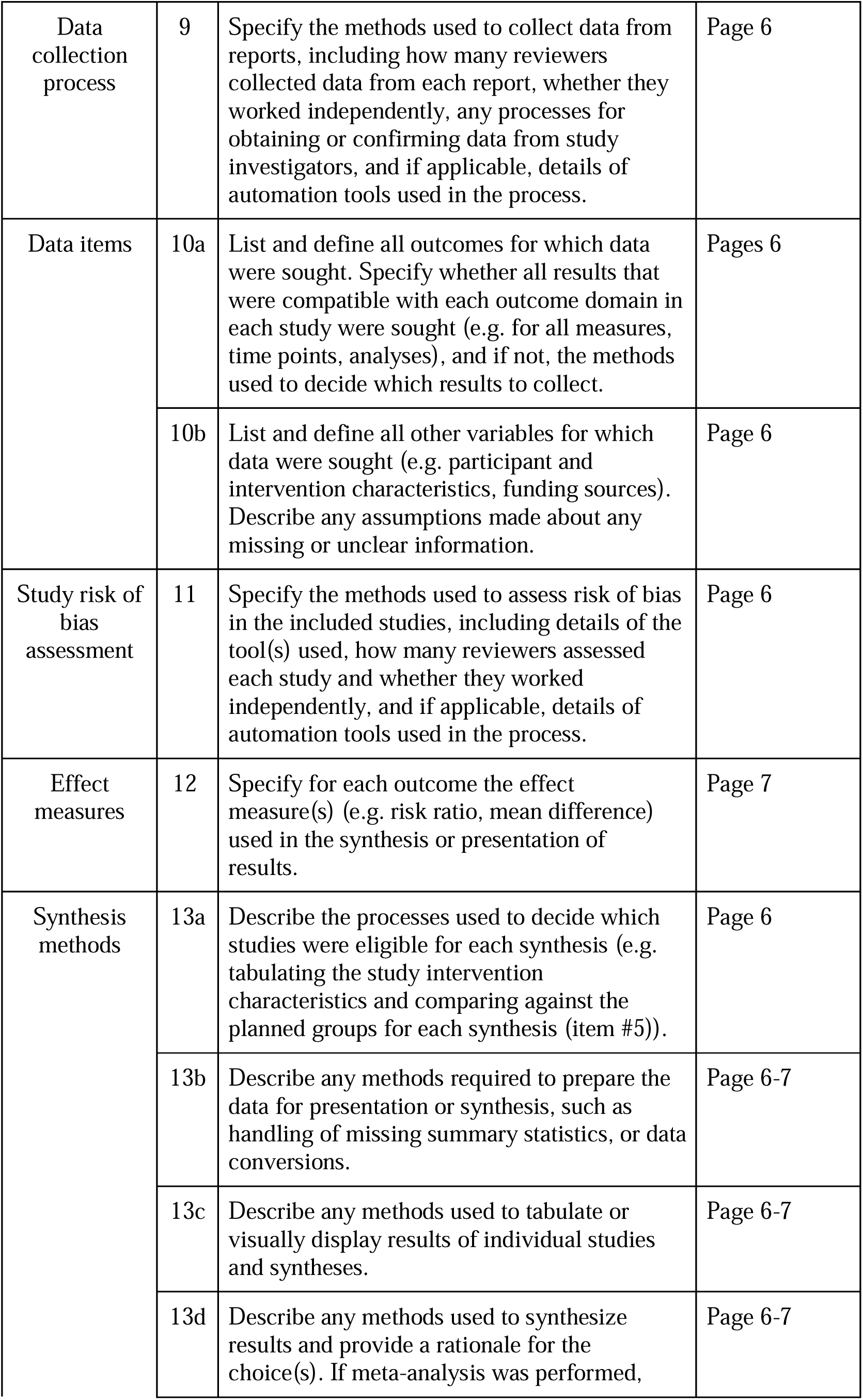
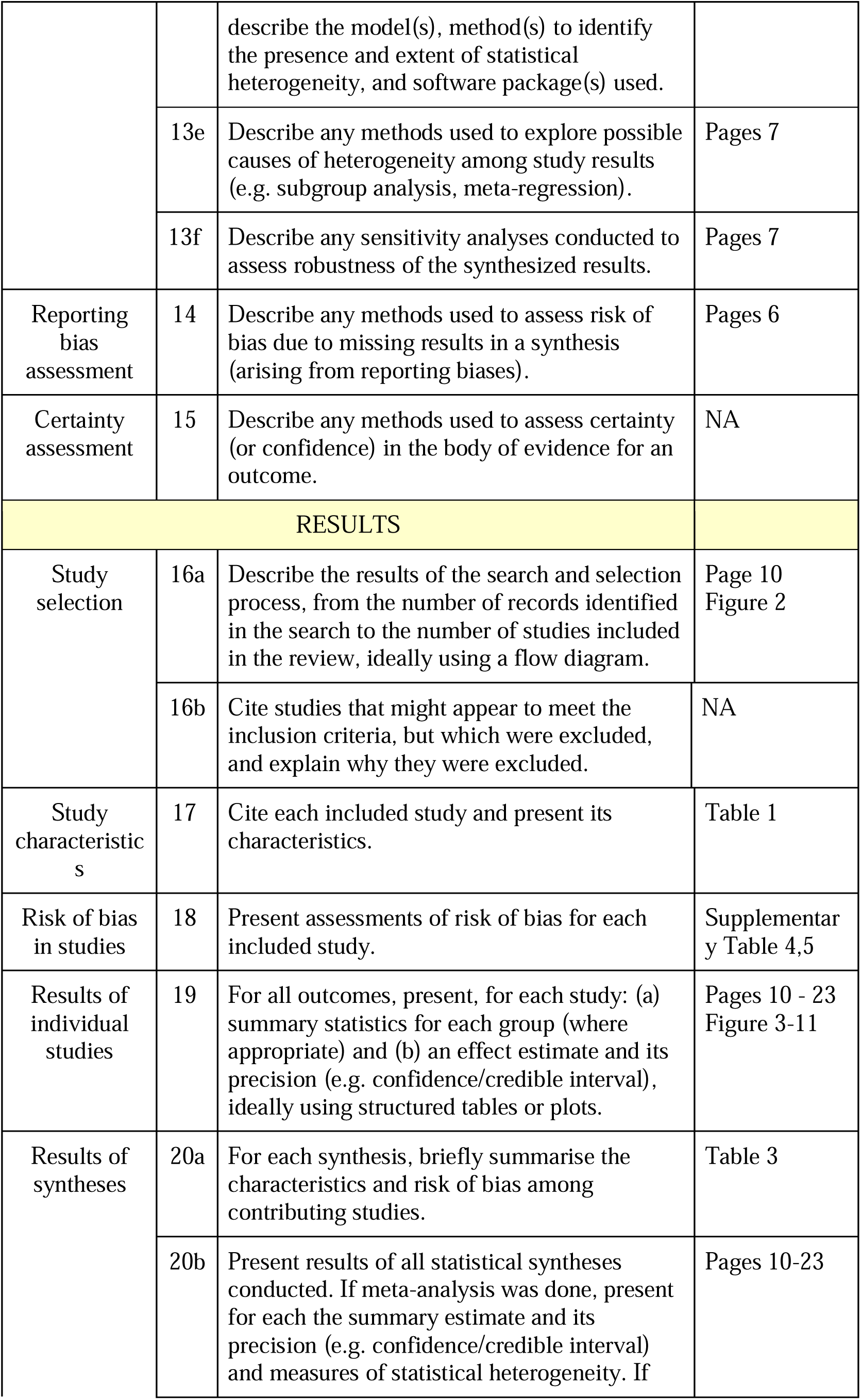
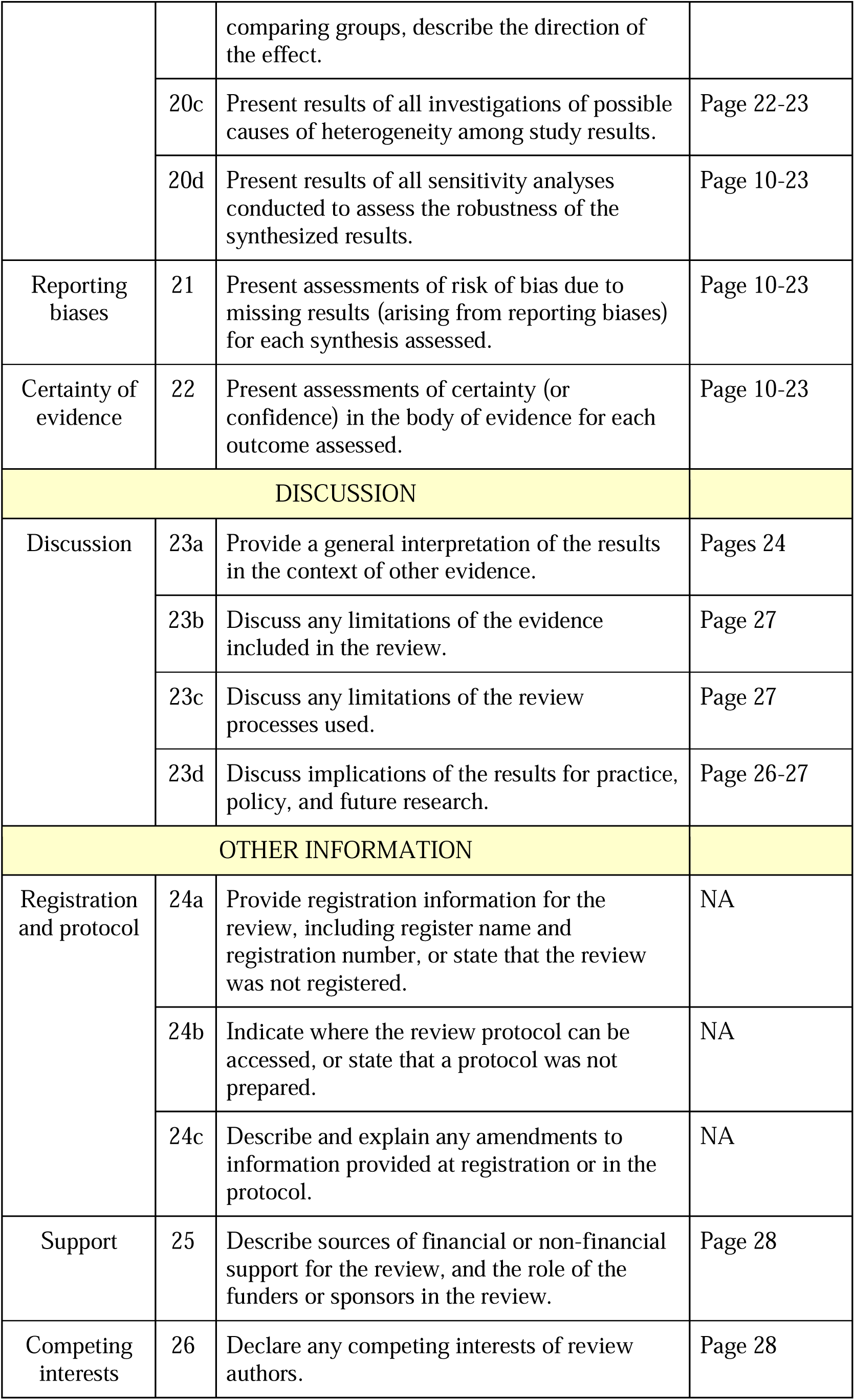
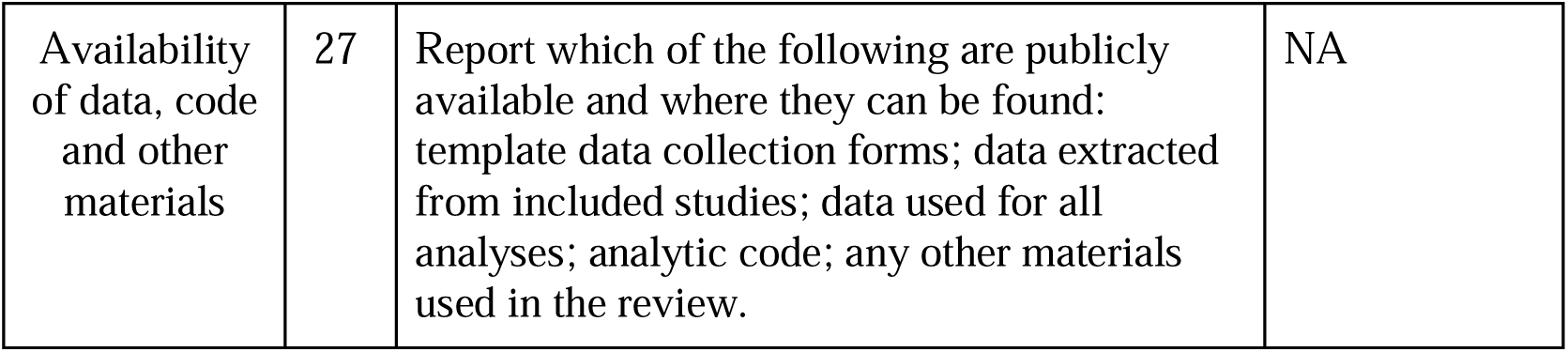
PRISMA 2020 Checklist.

**Supplementary Table 2.** PRISMA 2020 for Abstract Checklist.

| Section and Topic | Item # | Checklist item | Reported (Yes/No) |
| --- | --- | --- | --- |
| **TITLE** |  |  |  |
| Title | 1 | Identify the report as a systematic review. | Yes |
| **BACKGROUND** |  |  |  |
| Objectives | 2 | Provide an explicit statement of the main objective(s) or question(s) the review addresses. | Yes |
| **METHODS** |  |  |  |
| Eligibility criteria | 3 | Specify the inclusion and exclusion criteria for the review. | Yes |
| Information sources | 4 | Specify the information sources (e.g. databases, registers) used to identify studies and the date when each was last searched. | Yes |
| Risk of bias | 5 | Specify the methods used to assess risk of bias in the included studies. | Yes |
| Synthesis of results | 6 | Specify the methods used to present and synthesise results. | Yes |
| **RESULTS** |  |  |  |
| Included studies | 7 | Give the total number of included studies and participants and summarise relevant characteristics of studies. | Yes |
| Synthesis of results | 8 | Present results for main outcomes, preferably indicating the number of included studies and participants for each. If meta-analysis was done, report the summary estimate and confidence/credible interval. If comparing groups, indicate the direction of the effect (i.e. which group is favoured). | Yes |
| **DISCUSSION** |  |  |  |
| Limitations of evidence | 9 | Provide a brief summary of the limitations of the evidence included in the review (e.g. study risk of bias, inconsistency and imprecision). | Yes |
| Interpretation | 10 | Provide a general interpretation of the results and important implications. | Yes |
| **OTHER** |  |  |  |
| Funding | 11 | Specify the primary source of funding for the review. | Yes |
| Registration | 12 | Provide the register name and registration number. | Yes |

**Supplementary Table 3.**
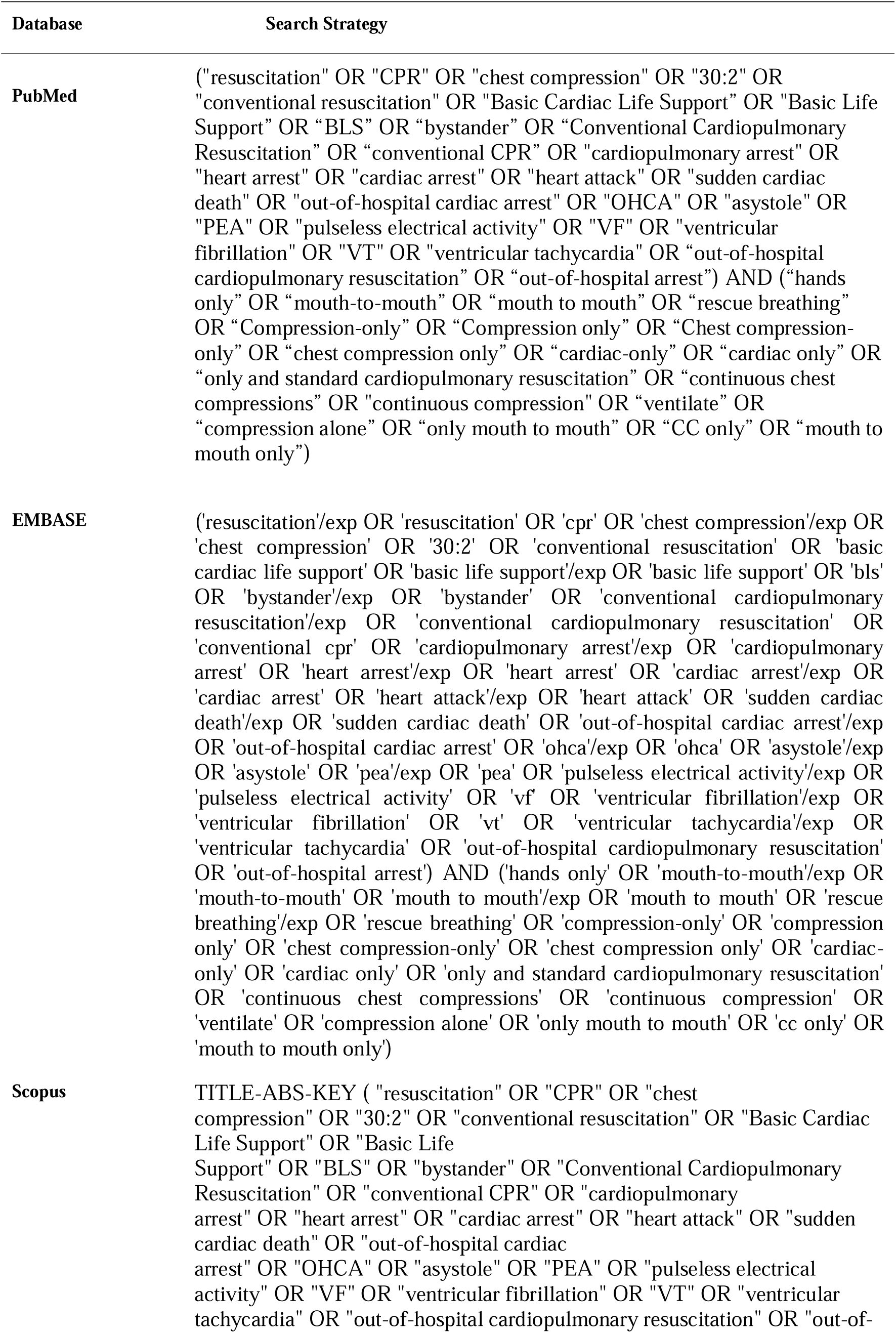
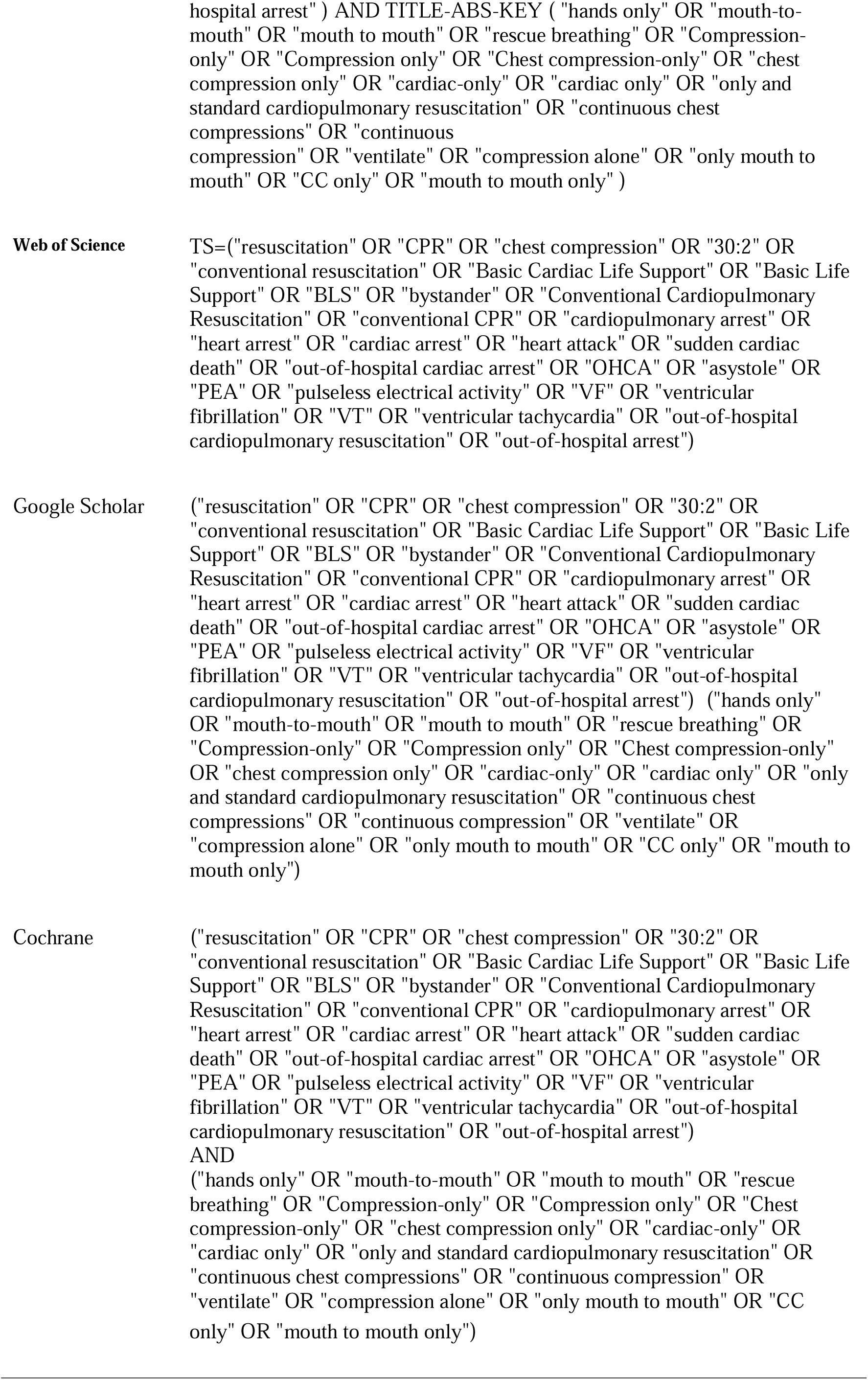
Search Strategies.

**Supplementary Table 4.** Quality assessment of the included observational studies using the Newcastle-Ottawa Scale.

| Author (ref.) | selection |  |  |  | comparability | outcome |  |  | Total score | Total Quality |
| --- | --- | --- | --- | --- | --- | --- | --- | --- | --- | --- |
|  | 1 | 2 | 3 | 4 | 1 | 1 | 2 | 3 |  |  |
| Bobrow et al. (1) | * | * | * | * | ** | * | * | * | 9 | Good |
| Bohm et al. (2) | * | * | * | * | * | * | * | * | 8 | Good |
| Iwami et al. (3) | * | * | * | * | * | * | * | * | 8 | Good |
| Javaudin et al. (4) | * | * | * | * | ** | * | * | * | 9 | Good |
| Kaneto et al. (5) | * | * | * | * | ** | * | * | * | 9 | Good |
| Kitamura et al. (6) | * | * | * | * | ** | * | * | * | 9 | Good |
| Olasveengen et al. (7) | * | * | * | * | ** | * | * | * | 9 | Good |
| Ong et al. (8) | * | * | * | * | ** | * | * |  | 8 | Good |
| Riva et al. (9) | * | * | * | * | ** | * | * | * | 9 | Good |
| Schmicker et al. (10) | * | * | * | * | ** | * | * | * | 9 | Good |
| SOS-KANTO study group (11) | * | * | * | * | ** | * | * | * | 9 | Good |
| Waalewijn et al. (12) | * | * | * | * | ** | * | * | * | 9 | Good |
| Wnent et al. (13) | * | * | * | * | ** | * | * | * | 9 | Good |

**Supplementary Table 5.**
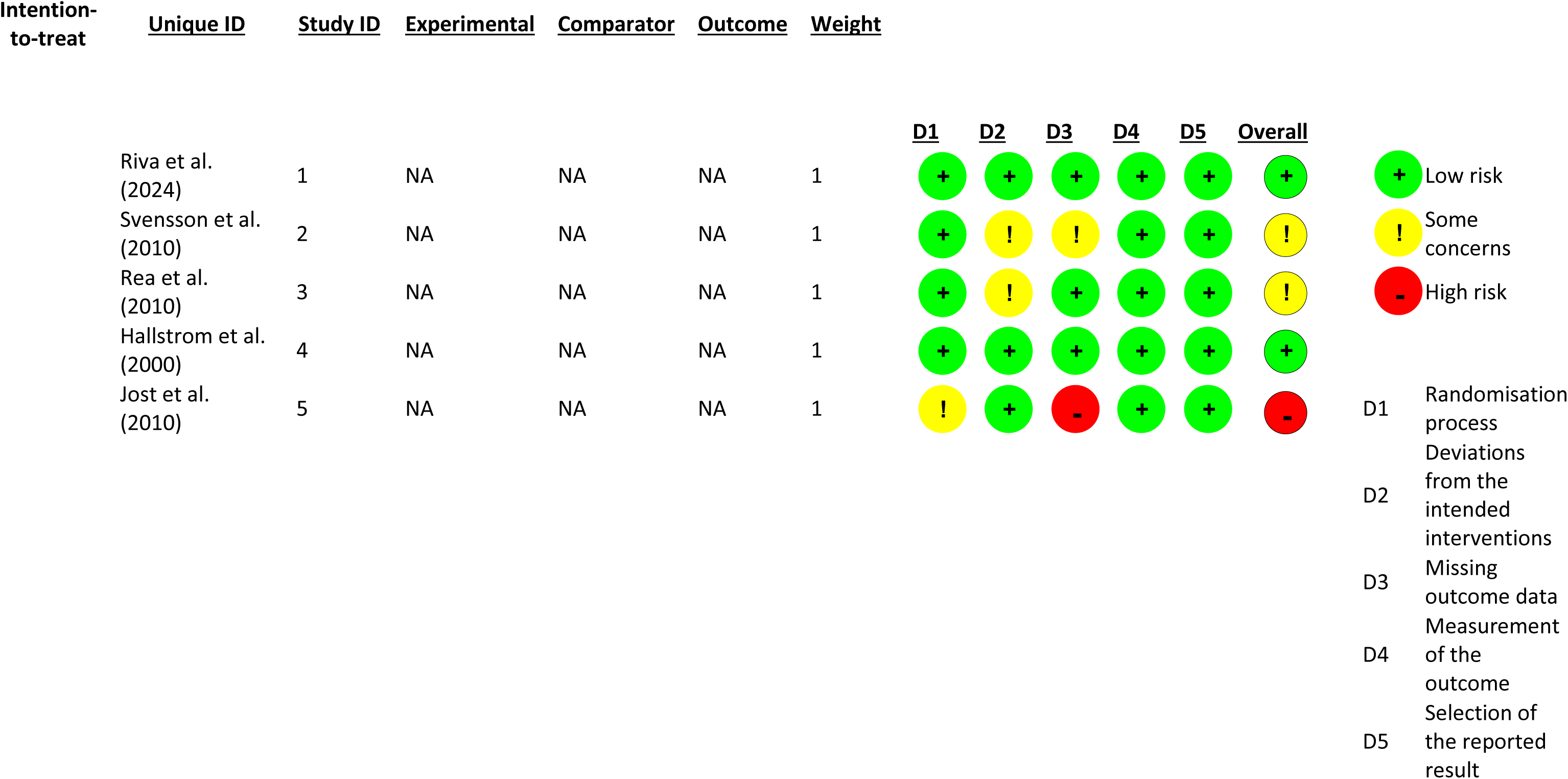
Quality assessment of the included RCTs studies using the ROB-2.

## Notes

### Competing Interest Statement

The authors have declared no competing interest.

### Author Declarations

Meta-analysis done from published data

